# Left ventricular myocardial molecular profile of human diabetic ischaemic cardiomyopathy

**DOI:** 10.1101/2024.12.06.24318627

**Authors:** Benjamin Hunter, Yunwei Zhang, Dylan Harney, Holly McEwen, Yen Chin Koay, Michael Pan, Cassandra Malecki, Jasmine Khor, Robert D. Hume, Giovanni Guglielmi, Alicia Walker, Shashwati Dutta, Vijay Rajagopal, Anthony Don, Mark Larance, John F. O’Sullivan, Jean Yang, Sean Lal

**Author notes:** Corresponding author: Sean Lal MD, PhD. Co-senior authors (John F. O’Sullivan, Jean Yang, Sean Lal).

## Abstract

Ischaemic cardiomyopathy is the most common cause of heart failure and often coexists with diabetes mellitus which worsens patient symptom burden and outcomes. Yet, their combined effects are seldom investigated and are poorly understood. To uncover the influencing molecular signature defining ischaemic cardiomyopathy with diabetes, we performed multi-omic analyses of ischaemic and non-ischaemic cardiomyopathy with and without diabetes against healthy age-matched donors. Tissue was sourced from pre-mortem human left ventricular myocardium. Fatty acid transport and oxidation proteins were most down-regulated in ischaemic cardiomyopathy with diabetes relative to donors. However, the down-regulation of acylcarnitines, perilipin, and ketone body, amino acid and glucose metabolising proteins indicated lipid metabolism may not be entirely impaired. Oxidative phosphorylation, oxidative stress, myofibrosis, and cardiomyocyte cytoarchitecture also appeared exacerbated principally in ischaemic cardiomyopathy with diabetes. These findings indicate diabetes confounds the pathological phenotype in heart failure, and the need for a paradigm shift regarding lipid metabolism.

## Introduction

Heart failure (HF) is a global epidemic and leading cause of death affecting 1-2% of adults in developed countries, with the most common causes being ischaemic cardiomyopathy (ICM) and non-ischaemic (dilated) cardiomyopathy (NICM)^1–3^. ICM is an acquired cardiomyopathy typically due to coronary artery disease (CAD) which primarily affects the left ventricle (LV) ^4,5^. NICM can involve both ventricles and is considered to arise from a mix of acquired and genetic etiologies^6–8^.

ICM and NICM both express ventricular dilatation and dysfunction and are the most common causes for heart transplantation^2,5,6,9^. Yet, ICM transplant recipients have an elevated risk of acute cardiac events and a reduced 10-year survival rate (50%) compared to NICM (84%), often due to underlying risk factors promoting CAD progression, including hypertension and metabolic syndrome^10^. These risk factors also contribute to the development of type 2 diabetes mellitus (DM), which is often concomitant with ICM in HF patients^11–13^. DM, in turn, is associated with dysregulation of fatty acid oxidation (FAO) pathways, with fatty acids being a principal energy source for the heart^14–18^. These metabolic processes, in addition to the extracellular matrix (ECM) and contractile complexes of the myocardium, have not been comprehensively investigated in human hearts, with ICM and DM seldom investigated together but frequently studied in isolation^19,20^. Given that DM exacerbates HF symptoms and increases mortality in HF^21,22^, there is a need to understand the impact of DM on HF.

Therefore, the aim of this study was to perform an extensive multi-omic analyses on cryopreserved pre-mortem human left ventricular myocardium to uncover the influence of DM in end-stage HF and to delineate the molecular characteristics of ICM with DM (ICM-DM), thereby creating a comprehensive molecular resource of DM-related-HF.

## Results

Myocardial multi-omic mass spectrometry (MS) differential analyses were performed on all end-stage HF conditions vs healthy age-matched donors (AMD) wherein DM and no DM groups were contrasted against each other (Fig. 1). Demographic data can be seen in Supplementary Table 1. Multidimensional scaling (MDS) plots were used to visualise individual and group similarity via a distance matrix for each MS analysis (Supplementary Fig. 1).

**Figure 1.**
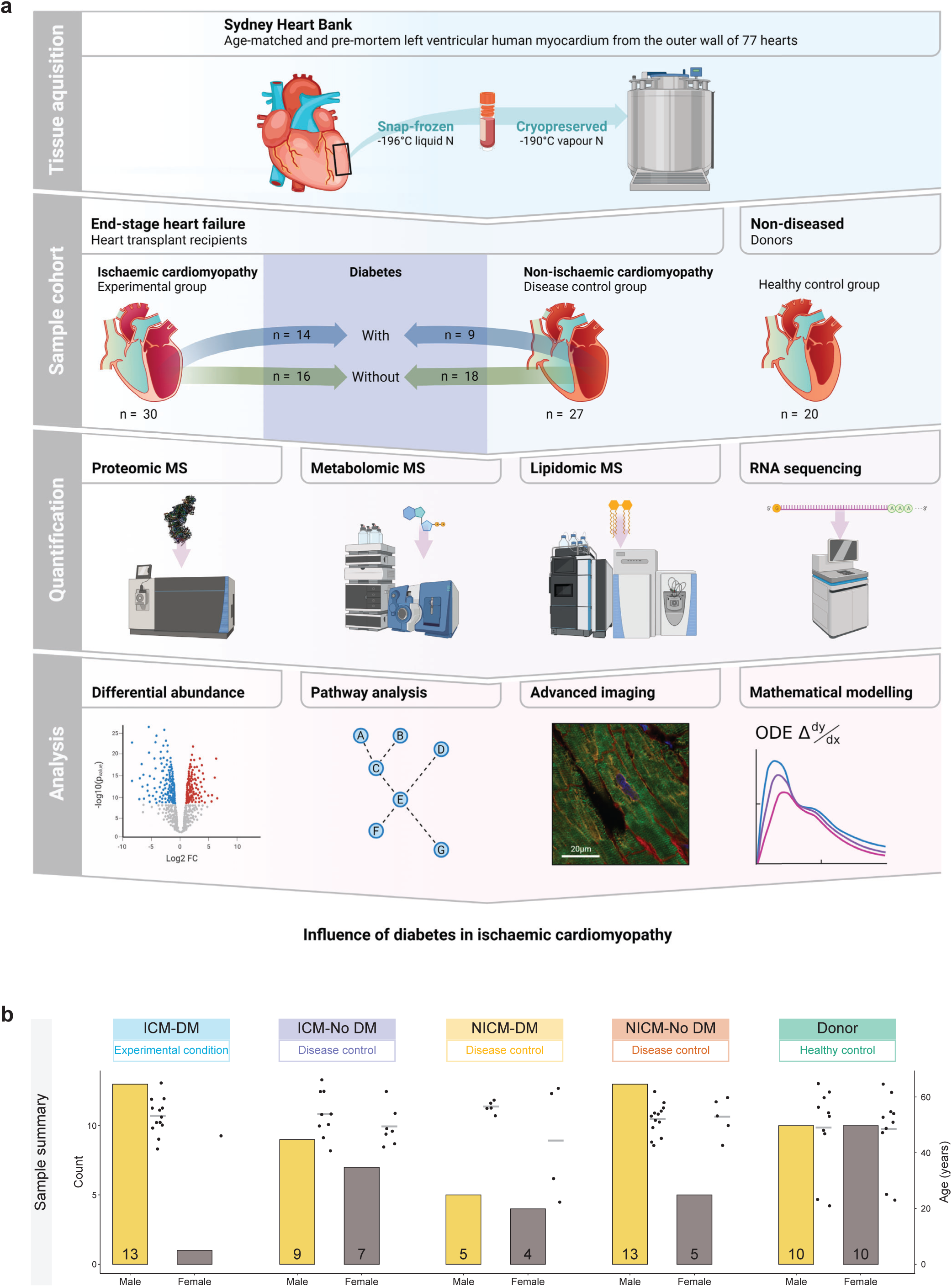
Overview of experimental design, workflow summary and sample summary. **a**, Workflow overview. Created with BioRender.com. **b**, Sample summary displaying sex and age of conditions.

### Differential molecular analysis of ischaemic cardiomyopathy with diabetes

Analyses of HF conditions with and without diabetes compared against AMD can be found in Supplementary Fig. 2 and Supplementary Tables 2-7. When focused on individual HF conditions vs AMD, proteomic analysis resulted in 593 down-regulated and 654 up-regulated proteins in ICM-DM of which, 1098 were only differentially abundant (DA) in the ICM-DM group of ICM such as regulator of microtubule dynamics (RMDN3), α-actinin 1 (ACTN1), mitochondrial isocitrate dehydrogenase (IDH2), and mitochondrial acetyl-CoA acetyltransferase (ACAT1) (Fig. 2a,d,g, Supplementary Tables 8,11). Cross-comparison of HF conditions vs AMD identified serum amyloid A1 (SAA1) was uniquely and greatly down-regulated in ICM-DM. Other highly DA proteins specific to ICM-DM included neogenin (NEO1), numerous NDUF complex I (CI) subunits such as NDUFS3 and muscle aldolase (ALDOA) (Supplementary Fig. 3a,d,g, Supplementary Tables 15,18). Proteins specifically DA in DM groups included multiple NDUFs, cysteine protease inhibitor FETUB, acyl-CoA oxidase (ACOX2), mitochondrial acetyl-CoA acyltransferase (ACAA2), and extracellular matrix (ECM)-related cytoskeletal protein talin 1 (TLN1). Some of the most significant ICM-DM proteins were also shared with the other HF groups including complement factor D (CFD), glutamic--pyruvic transaminase 2 (GPT), and mitochondrial acyl-CoA synthetase (ACSS3).

**Figure 2.**
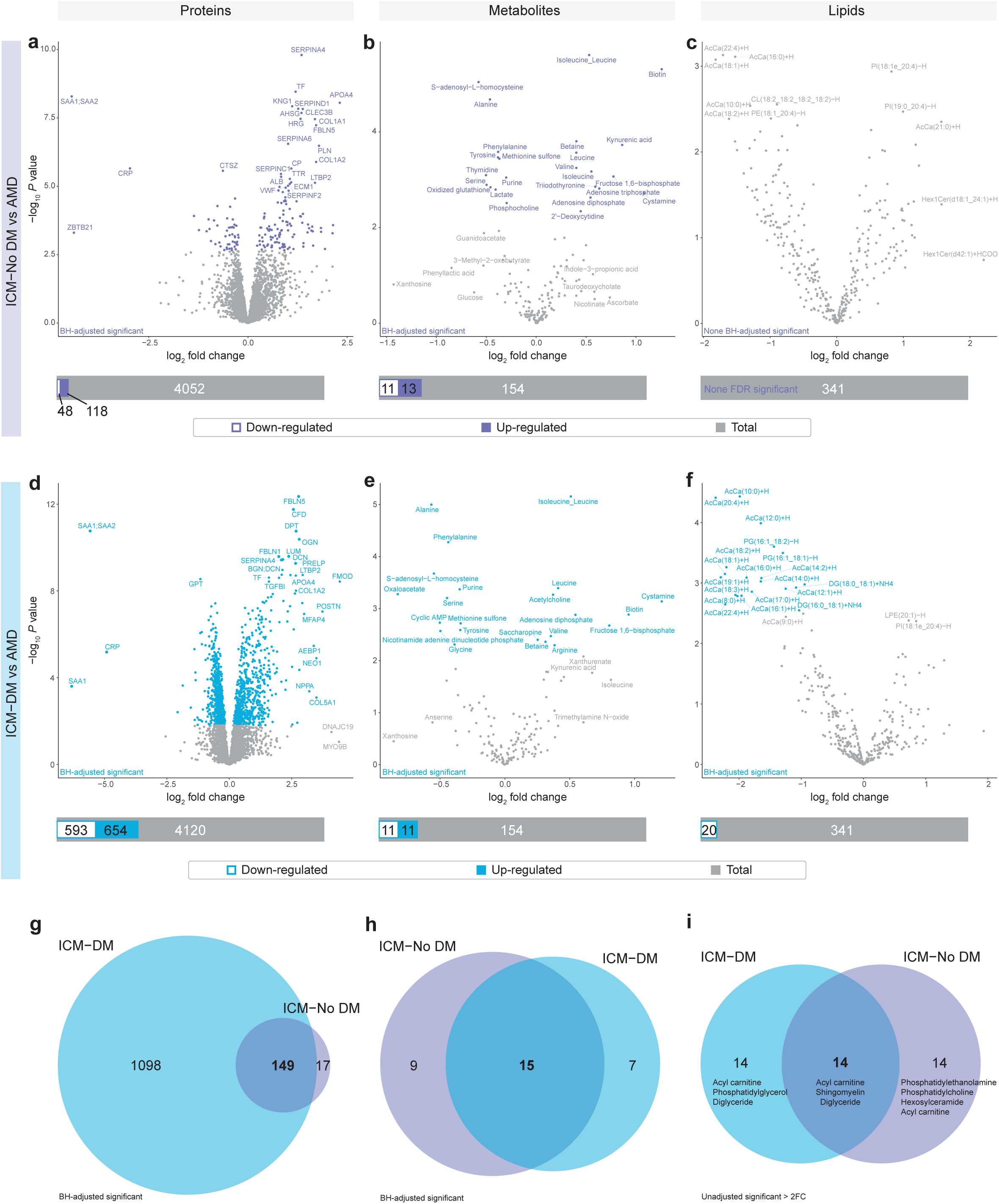
Human left ventricular myocardial differential analysis in protein, metabolite, and lipid abundance between ischaemic cardiomyopathy with (ICM-DM) and without diabetes (ICM-No DM) and age-matched donors (AMD). **a-f**, Differential abundance was determined following Benjamini-Hochberg false discovery rate adjustment (FDR) of *P* values (FDR < 0.05). Superimposed bar plots summarise the number of significantly down-regulated (white-filled bar) and up-regulated (colour-filled bar) molecules relative to the total number of molecules analysed (grey bar). **a**-**c**, ICM-No DM vs AMD. **d**-**f**, ICM-DM vs AMD. **g**-**h**, ICM-DM and ICM-No DM vs AMD FDR significant differentially abundant proteins and metabolites. **i**, ICM-DM vs AMD and ICM-No DM vs AMD unadjusted significant (*P <* 0.05) lipids which were also > ±2-fold change (FC, in either direction). Lipid classes annotated in order of the number of lipids which were unadjusted significant.

In the metabolomic analysis, there were 11 down-regulated and 11 up-regulated metabolites in ICM-DM vs AMD of which 7 were DA in ICM-DM only in the ICM group. Oxaloacetate, acetylcholine, nicotinamide adenine dinucleotide phosphate, glycine, arginine and cyclic adenosine monophosphate (cAMP), a second messenger for intracellular signalling, were among the metabolites uniquely DA in ICM-DM relative to the other forms of HF (Fig. 2b,e,h, Supplementary Figs. 3b,e,h, 5d-e, Supplementary Tables 9,13,16,19). Metabolites which were uniquely up-regulated in ICM included fructose 1,6-bisphosphate and adenosine diphosphate (ADP). Alanine (down-regulated) and isoleucine/leucine amino acids (up-regulated) were found to be commonly DA in all HF conditions in the same direction. Cystamine featured the greatest FCs and was up-regulated in ICM-DM, ICM-No DM, and NICM-No DM. NAD^+^/NADH ratio was up-regulated in ICM-DM, ICM-No DM, and NICM-No DM vs AMD. NADP^+^/NADPH ratio was down-regulated in ICM-DM, ICM-No DM, and NICM-No DM.

Lipidomic analysis identified 20 down-regulated lipids in ICM-DM, 15 of which were medium-chain (4), long-chain (10), and very long-chain (1) acylcarnitines, as well as diacylglyerols (2) and phosphatidylglycerols (2), with no other HF condition presenting FDR significant lipid differential abundance individually (Fig. 2c,f, Supplementary Figs. 3c,f, 5f, Supplementary Tables 10,14,17,20).

To allow exploration of these differential analyses, we produced an interactive resource (shiny app, http://shiny.maths.usyd.edu.au/Human-Heart-ALL/).

### Enriched metabolic pathways and gene sets

To investigate implicated pathways, Gene Set Enrichment Analysis (GSEA) protein enrichment analyses via clusterProfiler were performed on the normalised data using targeted Kyoto Encyclopedia of Genes and Genomes (KEGG) and MitoCarta3.0 databases (Figs. 3a, Supplementary Fig. 4a, Supplementary Tables 21-24). Selected enriched mitochondrial/energetics associated pathways/gene sets were annotated, ranked, and cross-HF group comparisons were performed (Figs. 3c,d,g, Supplementary Fig. 4c,d).

**Figure 3.**
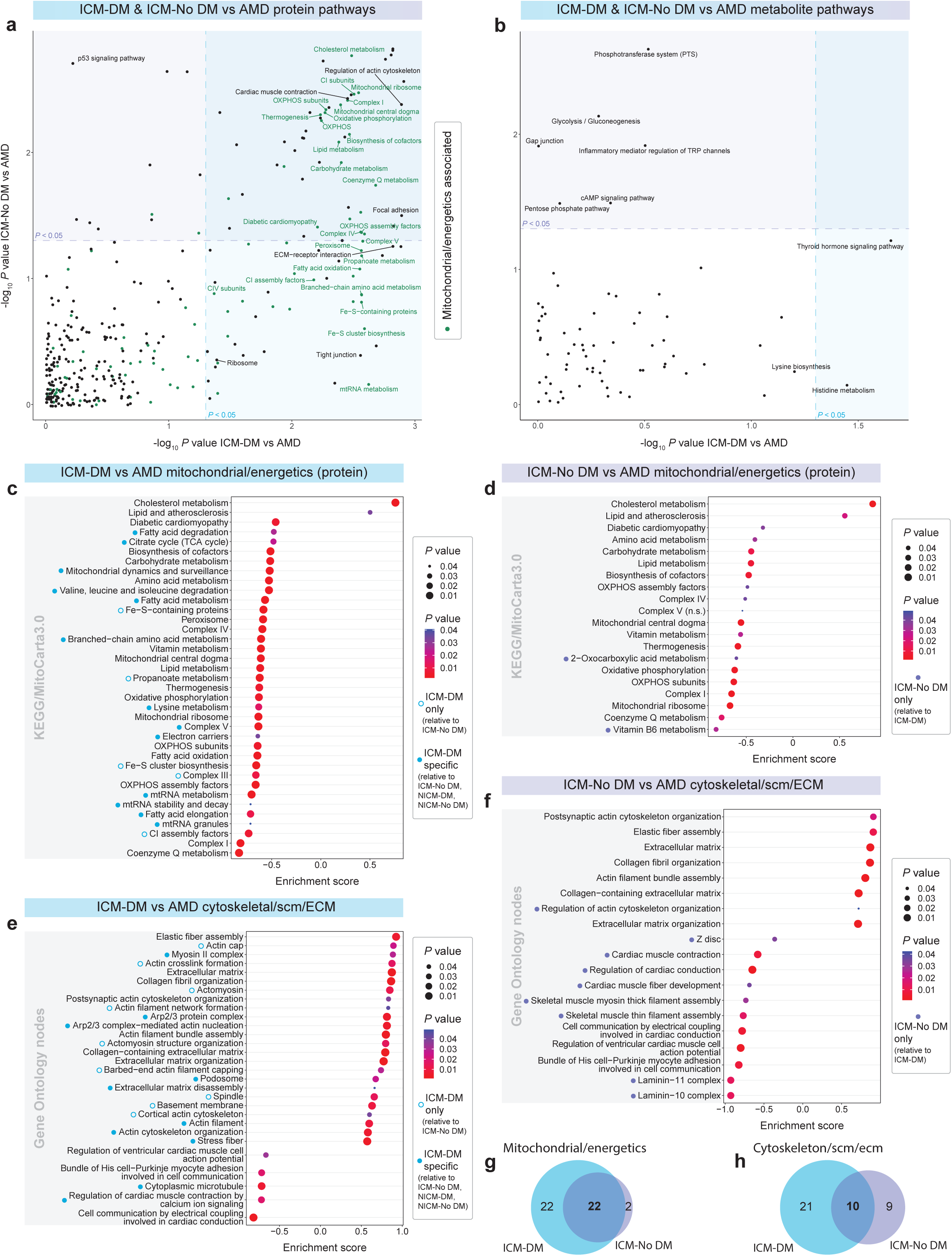
Human ischaemic cardiomyopathy with (ICM-DM) and without diabetes (ICM-No DM) vs healthy age-matched donor (AMD) protein and metabolite pathway analyses. **a**-**b**, Scatter plots of -log_10_ *P* value enriched KEGG and MitoCarta3.0 protein pathways/gene sets of ICM-DM vs AMD and ICM-No DM vs AMD where mitochondrial and energetics related pathways are coloured in green and significant (*P* < 0.05). Pathways in overlapping coloured regions were significant in both ICM-DM and ICM-No DM vs AMD. **a**, Enriched pathways from the proteomic mass spectrometry (MS) analysis. **b**, Enriched pathways from the metabolomic MS analysis. **c**-**f**, Gene Set Enrichment Analysis (GSEA) enrichment bubble plots of top ranked significantly enriched (*P* < 0.05) KEGG/MitoCarta3.0 pathways/gene sets and Gene Ontology nodes, respectively. **c**-**d**, From Fig. 3a. **e**-**f**, Cytoskeletal, sarcomeric (scm), and extracellular matrix (ECM) enriched Gene Ontology Biological Process and Cellular Component nodes from selected parent nodes. **g**, Mitochondrial/energetics associated pathways from Fig. 3a. **h**, All enriched Gene Ontology nodes which produced Fig. 3e-f.

Lipid metabolism, atherosclerosis, and cholesterol metabolism pathways were positively enriched in HF. Cholesterol metabolism was specifically enriched in ICM-DM, ICM-No DM, and NICM-DM. This enrichment was largely attributed to the up-regulation of intramyocardial chylomicron apolipoproteins, including APOA4, APOB, and APOC3, in all HF conditions, with all but one (APOL6) being up-regulated in ICM-DM. There were many common negatively enriched pathways/gene sets such as lipid metabolism, carbohydrate metabolism, thermogenesis, and diabetic cardiomyopathy in all HF groups. Additional pathways/gene sets were negatively enriched only in ICM-DM, such as the citrate cycle, mitochondrial RNA, fatty acid, branched-chain amino acid (BCAA), and lysine metabolism. Another notable negatively enriched metabolic pathway was propanoate metabolism, along with the peroxisome gene set, which were shared with NICM.

Oxidative phosphorylation (OXPHOS) and many of its subdivided gene sets were also observed as negatively enriched in all HF conditions, however, showed greater enrichment in ICM-DM wherein a greater number of the contained gene/proteins were FDR significant from the differential analysis (90 OXPHOS genes DA in ICM-DM out of 169; NICM-No DM, 35; NICM-DM, 27). All were down-regulated except for the up-regulation of HIG1 hypoxia inducible domain family member 1A (HIGD1A). CI subunits were most enriched in ICM-DM (enrichment score = -0.85) followed by NICM-DM (enrichment score = -0.76). Other proton pumping complexes and their subunit gene sets; CIII, CIV, and CV, were similarly negatively enriched in ICM-DM wherein CIII and CIV were co-enriched in NICM-DM and CV was ICM-DM specific. Gene sets mediating electron transport for OXPHOS, such as iron-sulfur cluster biosynthesis (enriched in DM) coenzyme Q metabolism (enriched in ICM), and electron carriers were also negatively enriched in ICM-DM.

The clusterProfiler KEGG enriched pathway analysis on the normalised metabolomic MS data identified a positive enrichment in histidine metabolism and a negative enrichment in thyroid hormone signalling pathways exclusive to ICM-DM vs AMD (Fig 3b, Supplementary Fig. 4b, Supplementary Tables 25-28).

### Extracellular matrix, cytoskeletal and sarcomeric enriched gene sets

GSEA Gene Ontology (GO) enrichment analysis was also performed on selected cytoskeletal, sarcomeric, and ECM parent nodes from the Biological Process (BP) and Cellular Component (CC) libraries vs AMD (Fig. 3e,f,h, Supplementary Fig. 4e,f, Supplementary Tables 29-32). Cardiomyocyte contraction and conduction-related nodes were seen to be negatively enriched specific to ICM conditions whereas regulation of cardiac muscle contraction by calcium ion signalling was unique to ICM-DM. This enrichment was largely attributable to the down-regulation of cardiac troponin I (TNNI3), sarcoplasmic reticulum Ca2^+^ ATPase (ATP2A2), Na^+^/Ca2^+^ exchanger (SLC8A1), ryanodine receptor (RYR2), and calsequestrin 2 (CASQ2) as determined from the differential analysis. ICM-DM-specific positive enrichment was observed in many actin/cytoskeletal nodes such as Arp2/3 protein complex, podosome, actin filament, and stress fibre nodes, where cytoplasmic microtubule (strongly influenced by SAA1, TPT1, and MTUS2) was negatively enriched. GO nodes only positively enriched in DM included barbed-end actin filament capping, actin cap, and actomyosin structure organization and actomyosin nodes wherein ICM-DM showed deviation from NICM-DM and No-DM in higher myosin heavy chain (MYH10, MYH11, MYH9, and MYL9) FCs. Multiple ECM nodes were positively co-enriched in all HF conditions, but more proteins were observed to be FDR up-regulated in ICM-DM from the differential analysis, such as in elastic fiber assembly and collagen fibril organization, strongly influenced by collagens (COLs), with unique up-regulation in ICM-DM in proteins such as FKBP prolyl isomerase 10 (FKBP10) and elastin microfibril interfacer 1 (EMILIN1). We previously reported an increased elastic fiber assembly and elastogenesis within both rat and human ICM^23^. Cytoskeletal node, actin filament assembly, was also positively co-enriched in HF.

Supplementary HF and ICM-DM GO enrichment analyses via PANTHER from significantly DA proteins vs AMD supported the above clusterProfiler enrichment results (Supplementary Tables 33-36, Supplementary Figs. 5a-c, 6a-b).

### Mitochondria and oxidative phosphorylation simulated modelling

To study the potential physiological consequences of reduced CI activity in ICM-DM (Fig. 4a), we employed a biophysical model of OXPHOS (Fig. 4b-d). The model was validated using rat cardiomyocyte OXPHOS data due to its availability over human data. Given the metabolic differences between rat and human cardiac myocytes^24^, the model could not make quantitative predictions but was used to infer trends in physiological variables related to decreased CI activity. The simulations showed that decreased CI activity alone has little effect on the function of OXPHOS, which is consistent with results in prior theoretical studies^25^, and experimental studies in animals^26,27^. Figure 4e depicts an intra-mitochondrial schematic representing all DA proteins in ICM-DM vs AMD directly involved in OXPHOS.

**Figure 4.**
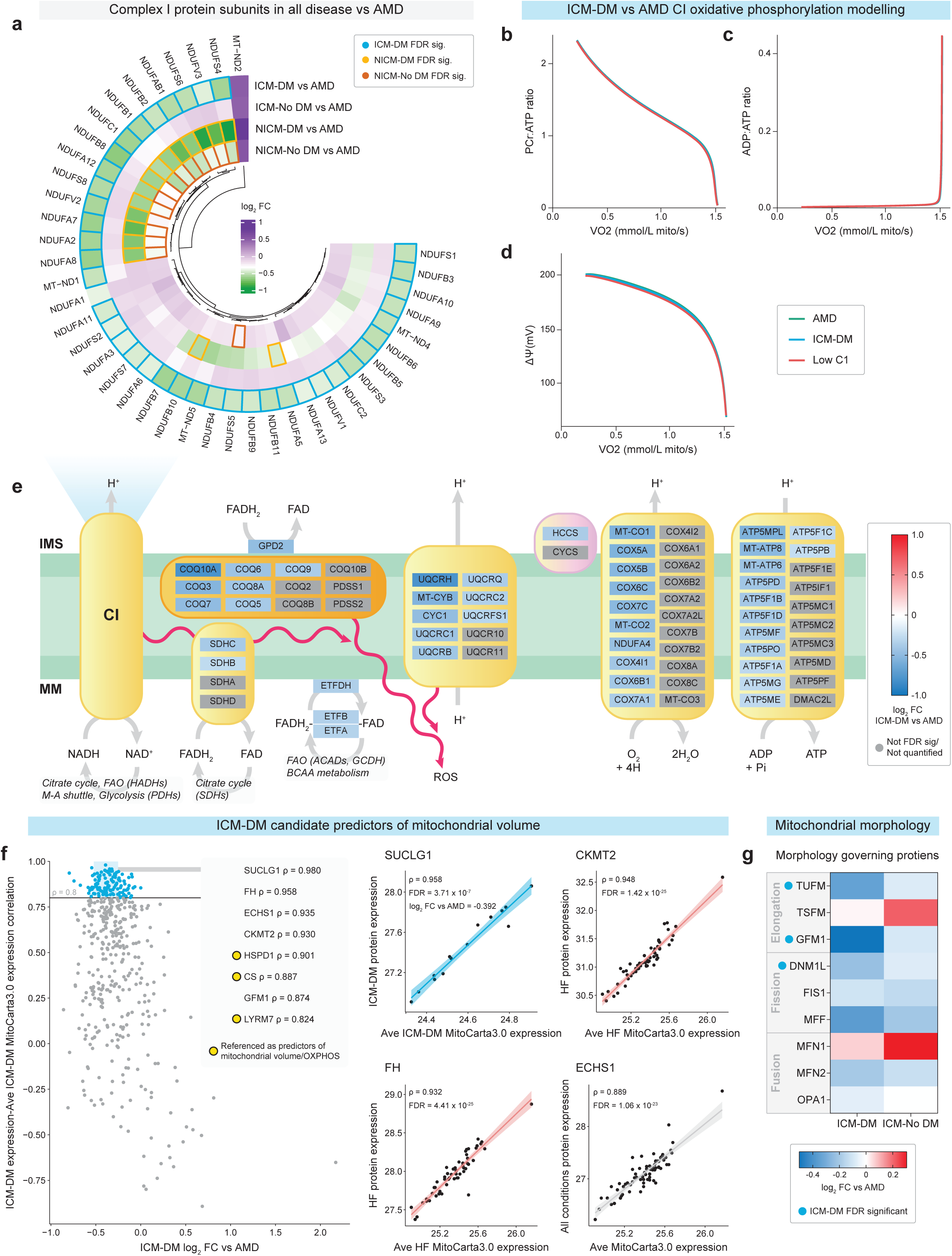
Ischaemic cardiomyopathy with diabetes (ICM-DM) influence on oxidative phosphorylation, mitochondrial composition and mitochondrial morphology. **a**, Quantified electron transport chain complex I (CI) subunits from proteomic mass spectrometry (MS) showing the log_2_ fold change (FC) of all heart failure (HF) conditions; ICM-DM, ICM without diabetes (ICM-No DM), non-ischaemic cardiomyopathy with diabetes (NICM-DM), and NICM without diabetes (NICM-No DM), vs age-matched donors (AMD). Significance was determined after Benjamini-Hochberg false discovery rate adjustment (FDR) of *P* values (FDR < 0.05). **b**-**d**, Simulations of the computational model of oxidative phosphorylation. The groups correspond to AMD myocytes (green; baseline CI activity), ICM-DM myocytes (blue; complex I activity −29.4%, based on proteomic MS) and myocytes with even lower CI expression (red; CI activity −40%). **d**, mitochondrial membrane potential (ΔΨ) are plotted against the oxygen consumption rate VO2. **e**, Schematic of oxidative phosphorylation depicting FDR significant proteins where colour represents log_2_ FC in ICM-DM vs AMD. Proteins not FDR significant or not quantified/analysed represented in grey. CI to CV; left to right. IMS, intermembrane space; MM, mitochondrial matrix; FAO, fatty acid oxidation; ROS, reactive oxygen species. **f**, Log_2_ FC of ICM-DM mitochondrial proteome vs AMD and correlation of ICM-DM individual mitochondrial protein expression to the average relative expression of the ICM-DM mitochondrial proteome across each patient. Only proteins quantified in all samples were analysed. **g**, Quantified proteins influencing mitochondrial morphology.

Leveraging the opportunity of analysing the pre-mortem human myocardium in healthy and end-stage HF conditions, we sought the protein with which expression per individual best correlated with the combined average expression of the mitochondrial proteome (MitoCarta3.0) as a candidate for predicting mitochondrial volume in ICM-DM, HF, or any condition. Succinate-CoA ligase SUCLG1 was the strongest predictor of ICM-DM mitochondrial volume (ρ = 0.958), while mitochondrial creatine kinase (CKMT2) (ρ = 0.948) and fumarate hydratase (FH) (ρ = 0.932) were best for HF. Enoyl-CoA hydratase ECHS1 (ρ = 0.889) was most suitable across all conditions (Fig. 4f, Supplementary Tables 37-39). These were alongside previously referenced proteins as predictors of mitochondrial volume/OXPHOS; citrate synthase (CS)^28,29^, heatshock protein HSPD1^30^, and LYR motif-containing protein 1 (LYRM1)^31^. Investigation of proteins regulating mitochondrial morphology identified the down-regulation of elongation factors G1 (GFM1) and Tu (TUFM), and fission protein dynamin 1 like (DNM1L) in ICM-DM (Fig. 4g).

As the ICM-DM group had a higher BMI (30.8 ± 5.1) than the other conditions, a BMI-average molecule abundance Pearson correlation test was also performed to identify a potentially significant contributary influence to the results from a high BMI/obesity, whereby, no significance was identified (Supplementary Tables 40,41,42, Supplementary Fig. 6c-e).

### RNA sequencing analysis, and RNA-protein correlation and co-regulation

To investigate whether significantly DA proteins may have been regulated at the transcription or post-translational level, or to identify differentially expressed non-coding sequences which could regulate protein expression, RNA sequencing (RNA-seq) was performed on selected ICM-DM (n = 7), ICM-No DM (n = 7) and AMD (n = 7) left ventricular myocardial tissues (Supplementary Table 43). It was found that 120 transcripts were down-regulated and 1,307 were up-regulated in ICM-DM vs AMD. Of these, 334 were differentially expressed in both ICM-DM and ICM-No DM. The 1,093 transcripts which were only differentially expressed in ICM-DM showed significant enrichment in the extracellular matrix GO CC node (FDR = 7.80 x 10^-17^) and the ECM-receptor interaction KEGG pathway (FDR = 0.0233) via STRING analysis (Supplementary Fig. 7a-c, Supplementary Tables 44,46).

Paired RNA and protein log_2_ FC of ICM groups were plotted to compare FC similarities compared to AMD (Fig. 5a,b). An RNA-protein Pearson correlation analysis was performed in each condition wherein 179 RNA transcripts and proteins were correlated only in ICM-DM, and nestin (NES), a biomarker for mitosis, was the only gene symbol co-correlated in all conditions (Fig.5c, Supplementary Tables 48-50). NES protein was up-regulated in all HF conditions, with the greatest FCs in DM, and was RNA-protein co-regulated in ICM-DM.

**Figure 5.**
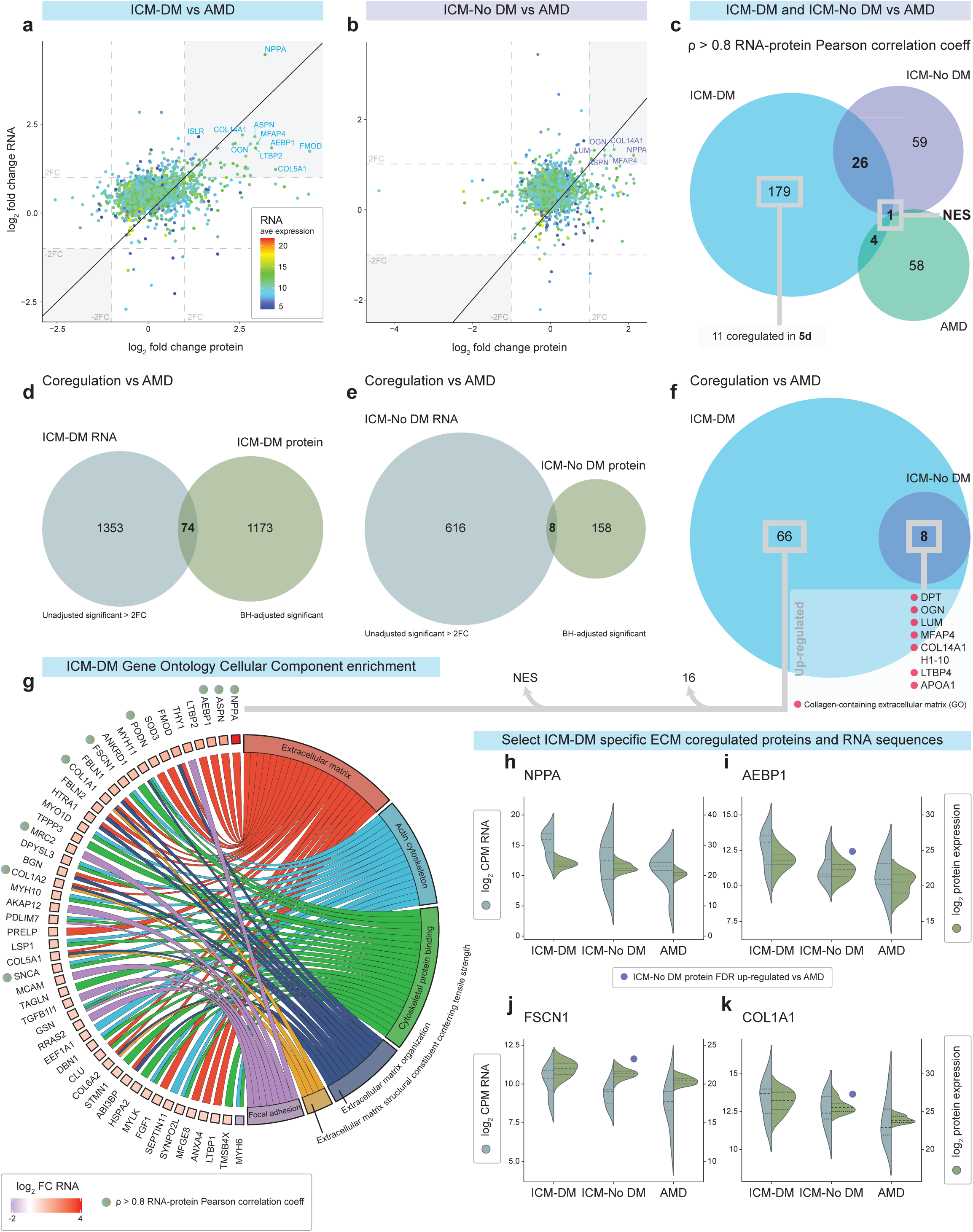
Ischaemic cardiomyopathy with (ICM-DM) and without diabetes (ICM-No DM) RNA-protein correlation and co-regulation vs age-matched donors (AMD). **a**, RNA and protein log_2_ fold change (FC) plot of ICM-DM vs AMD. ICM-DM average RNA expression represented on a colour scale. **b**, RNA and protein log_2_ FC plot of ICM-No DM vs AMD. **c**, Gene symbols which had an RNA-protein Pearson correlation coefficient of ρ > 0.8 in ICM-DM, ICM-No DM and AMD. **d**, ICM-DM vs AMD significant differentially expressed RNA (*P* < 0.05 and > ±2-FC) and protein (FDR < 0.05) gene symbols. **e**, ICM-No DM vs AMD significant RNA and protein gene symbols. **f**, From Fig. 5d, 5e, including list of gene symbols RNA-protein co-regulated in both ICM-DM and ICM-No DM vs AMD. **g**, Gene Ontology Cellular Component enrichment chord plot of extracellular, cytoskeletal and focal adhesion related gene symbols from Fig. 5f which were co-regulated only in ICM-DM vs AMD with RNA log_2_ FC represented. RNA-protein Pearson correlation coefficient of ρ > 0.8 indicated. **h**-**k**, Selected gene symbols of interest from Fig. 5g showing log_2_ transformed RNA and protein expression. Purple circles highlight the gene symbols which were also ICM-No DM vs AMD significant differentially expressed (FDR < 0.05) in the proteomic analysis.

It was found that 74 gene symbols were RNA-protein co-regulated in ICM-DM vs AMD in the same direction, 66 of which were only observed in ICM, and all ICM-No DM co-regulated gene symbols were shared by ICM-DM (Fig. 5d-f, Supplementary Table 52). A GO chord plot of selected significantly enriched CC nodes was formed to summarise the RNA and protein co-regulated only in ICM-DM showing that 49 of the 66 gene symbols were contained within extracellular, cytoskeletal and focal adhesion gene sets (Fig. 5g). Of these, 9 gene symbols were also RNA-protein correlated such as natriuretic peptide A (NPPA), adipocyte enhancer-binding protein 1 (AEBP1), fascin (FSCN1), and COL1A2 (Fig. 5h-k).

ICM-DM and ICM-No DM Common DA gene symbols and GO nodes vs AMD we identified, as well as ICM-DM divergent (DA to AMD and ICM-No DM) protein and RNA transcripts (Supplementary Fig. 8, Supplementary Tables 12,45).

A sex analysis revealed no clustering, significant abundance or correlation of molecules despite the male bias in ICM-DM, however, AEBP1 protein abundance showed a positive trend in males (Supplementary Fig. 9, Supplementary Tables 55-58).

### Histochemistry

Histochemistry was performed to qualitatively observe the myocardial changes in ICM-DM described by proteomic MS. Diffuse interstitial and perivascular collagen fibril staining was observed in ICM vs comparable AMD myocardium, representing increased fibrosis (Fig. 6a-c). Macroscopic infarct/replacement fibrosis was avoided in all analyses (Supplementary Fig. 11a,b). Key proteins of interest were labelled and imaged via immunofluorescent confocal microscopy, with cardiomyocytes longitudinally oriented, based on down-regulated proteomic MS results and their biological/cited significance. ATP2A2/SERCA2 showed disruption/disorganisation within ICM-DM cardiomyocytes, particularly along the Z-discs, vs AMD, with TNNI3 co-localising along the I-band (Fig. 6d-f,m,n, Supplementary Figs. 10m, 11c,d). Increased interstitial space was noted between the sarcolemma of ICM-DM cardiomyocytes, with broader and disorganised staining of intramembrane glycoproteins indicating ECM linkage dysfunction. Disruption was also indicated in OXPHOS CI subunit NDUFS3 and mitochondrial elongation factor GFM1 proteins at the I-band (Fig. 6g-l,o,p. Supplementary Figs. 10m, 11e,f). Mitochondria were also labelled with Mito-ID where bright aggregates were attributed by the compounding mitochondrial OXPHOS-associated flavin autofluorescence with the highest concentrations in the perinuclear region signifying a notable reduction of both in ICM-DM. Majority of the flavin at this emission wavelength (500-550nm) was assumed to be in the form of free flavin adenine dinucleotide (FAD), followed by flavin-bound flavoproteins (ETFs) with the least attributed to acyl-CoA dehydrogenases (Supplementary Fig. 10a-d,k)^32–37^. ICM-DM T-tubules between mitochondria presented disorganisation at Z-disc locations. Glycolytic muscle aldolase (ALDOA), which binds to F-actin of the sarcomere and regulates cardiac hypertrophy^38,39^, was regularly distributed across the Z-disc in AMD but appeared dislocated in ICM-DM (Supplementary Figs. 10e-j,l, 11g,h).

**Figure 6.**
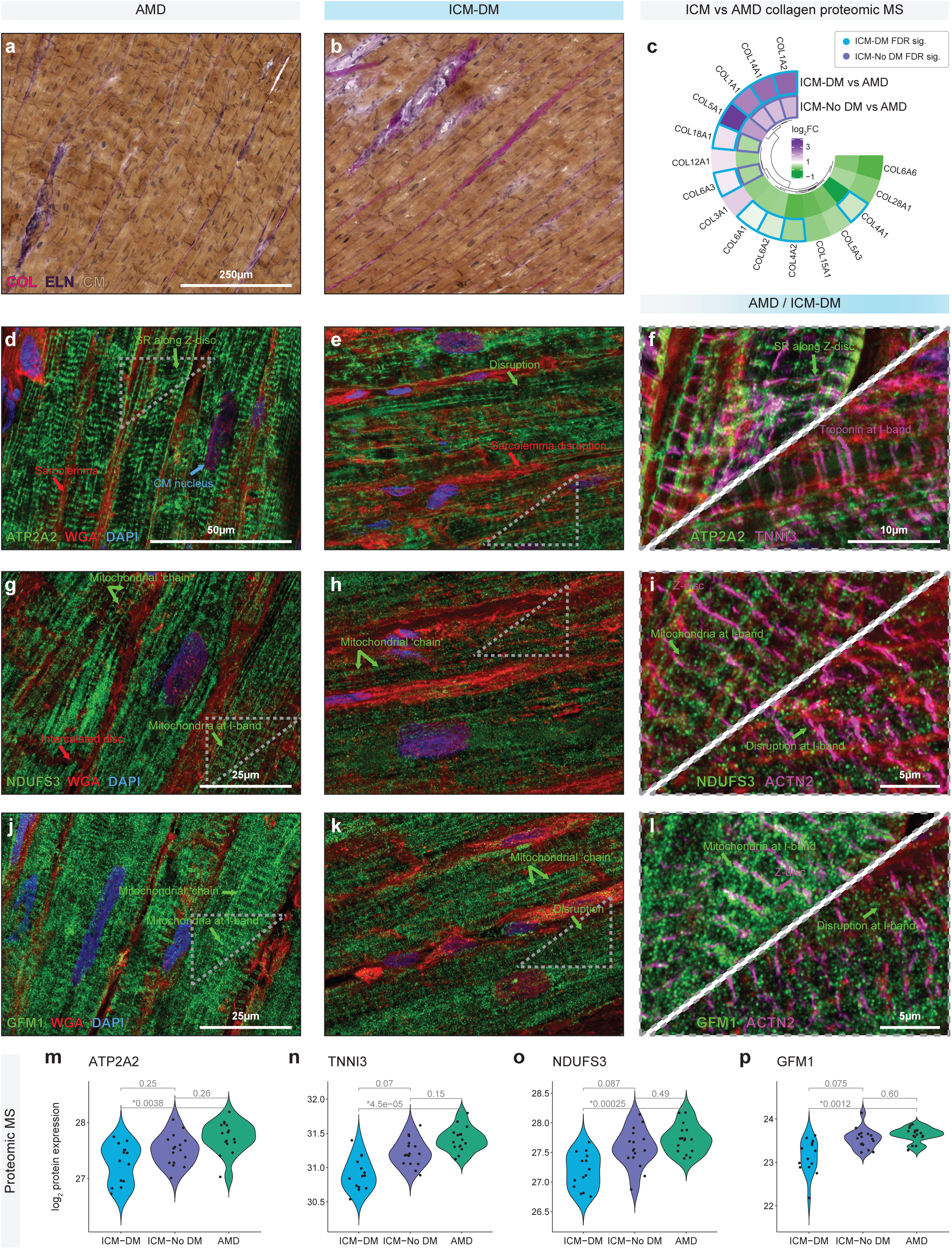
Histochemical visualisation of select key proteins in cryopreserved human left ventricular myocardium of ischaemic cardiomyopathy with diabetes (ICM-DM) and healthy age-matched donor (AMD) tissues. **a**-**b**, Verhoeff-Van Gieson stain of an AMD and ICM-DM with collagens (COL, magenta), elastin (ELN, purple), cardiomyocytes (CM, brown), and nuclei (blue) **c**, ICM-DM and ICM-No DM log_2_ fold change (FC) vs AMD quantified collagen proteins from proteomic mass spectrometry (MS) where Benjamini-Hochberg false discovery rate (FDR) adjusted significant differentially expressed proteins are indicated. **d**-**l**, Immunofluorescent confocal microscopy images depicting qualitative differences between AMD and ICM-DM in the labelled protein of interest (green) with a labelled reference protein (magenta). Membranes were stained with fluorophore-conjugated wheat germ agglutinin (WGA, red) and nuclei were stained using DAPI (blue). All images are 4.5µm thick Z-stacks, deconvolved using Huygens Professional, and compressed into a two-dimensional image using Fiji/ImageJ Maximum Intensity Projections. **m**-**p**, Proteomic MS log_2_ transformed quantification of immunohistological proteins of interest, ATP2A2, TNNI3, NDUFS3, and GFM1, in ICM-DM, ICM-No DM and AMD groups. FDR adjusted *P* values were (*, FDR < 0.05) between groups.

## Discussion

We have created a resource on the differential abundance of proteins, metabolites, lipids, and RNA in human left ventricular samples from end-stage ICM-DM compared to AMD and ICM-No DM, and in reference to NICM with and without DM.

Our results identified the up-regulation of circulating chylomicron apolipoproteins (APOs) in ICM-DM, suggesting that diabetes may drive reduced FAO. APOs are positively associated with high blood pressure and atherosclerosis and have been used as an indicator of metabolic syndrome and cardiac stress^40–43^. The down-regulation of components associated with the peroxisome proliferator-activated receptor (PPAR)-α pathway, of long-chain and very-long-chain fatty acid transporters (SLC27A1, CD36), cytosolic fatty acid binding proteins, acyl-CoA synthetases, and proteins involved in FAO, indicates a reduced utilisation of FA in ICM-DM. However, there was an up-regulation of albumin (ALB) and transmembrane medium-chain fatty acid transport (LRP1) in addition to a down-regulation of lipid droplet-surrounding perilipin (PLIN4) in ICM-DM, which may suggest an increased bioavailability of free fatty acids (FFAs) in the myocardium. Short-chain FAO may be unhindered or even up-regulated, as identified in a TAC-induced HF rat model^44^, as they do not require transmembrane or intracellular facilitatory transport.

It has been previously identified that ATP synthesis, of which FAO normally accounts for ∼50%-70% within the healthy myocardium, undergoes a metabolic shift to carbohydrate and ketone reliance in HF^16,45,46^. There is evidence that the heart prioritises glucose metabolism over FAO as a source of acetyl-CoA in the presence of oxidative stress or metabolic dysfunction^47–49^. Yet, paired with our proteomic results, medium-chain, long-chain, and very-long-chain acylcarnitines were down-regulated in HF vs AMD, particularly in ICM-DM, as well as diacylglycerol, an intermediate to triacylglycerol synthesis and lipid droplet formation, suggesting an increased fatty acid metabolism dependence or efflux/loss of fatty acids into the circulation in ICM-DM relative to other HF; used or lost. This concept of lipid efflux, supported by positive myocardial APOA1 RNA-protein co-regulation in ICM, to our knowledge, has not been described in the myocardium but been observed in chick skeletal muscle in response to lipid overload^50,51^.

Increased plasma acylcarnitines have been attributed as a characteristic of HF, including ICM-DM, and to identify high-risk patients^52–55^. Our results suggest down-regulation of FAO in HF as well as an association with reduced myocardial tissue acylcarntines, but there is a lack of consensus regarding whether FAO is down or up-regulated in HF and ICM-DM^45,56–59^. There was a down-regulation of PLIN4 in ICM-DM only, indicating the initiation of intracellular lipolysis and the release of FFAs for FAO. It has previously been shown that inactivation of PLIN4 results in the down-regulation of PLIN5 RNA and protein expression in the heart compared to other organs^60,61^. Reduced PLIN5 has been attributed to exaggeration of the HF phenotype, increased FAO and oxidative stress^62^, and has been considered a therapeutic target to prevent myocardial lipid accumulation^63^.

Myocardial lipogenesis and the resulting formation of lipid droplets have been a well-documented yet poorly understood feature of HF, ischaemia, insulin resistance and diabetes, obesity and metabolic syndrome^17,64–70^. This condition has been referred to as cardiac steatosis. We identified an overall reduced myocardial lipid content and to our knowledge, this process has not been investigated in human end-stage ICM-DM.

The end-stage ICM-DM heart showed signs of promoting glycolysis via intracellular glucose availability, as anticipated in HF and ischaemia due to pathological hypertrophy stimuli and oxidative stress^48,59,71,72^, by the up-regulation of glucose-trapping hexokinase (HK1, the first rate-limiting step of glycolysis), which was also observed in NICM in agreement with a recent study^14^.

Diabetes and insulin resistance have been shown to present with hyperglycaemia and impaired insulin-sensitive glucose transporter (SLC2A4/GLUT4) translocation to the plasma membrane and a reduced expression in the rat heart^18^. Our results also show the down-regulation of SLC2A4 in addition to an up-regulation of basal glucose transporters (SLC2A1/GLUT1) in ICM-DM with no DA in other forms of HF. This is in concordance with an early study in dogs^73^, however, an up-regulation of SLC2A1 was identified in rats with coronary ligation-induced HF^74^. As failing and ischaemic hearts have a greater dependence on glycolysis, impaired expression and translocation of SLC2A4 to the sarcolemma due to diabetes could result in an exacerbated reduction in systolic function^75^.

Ketone body, particularly 3-hydroxybutyrate, uptake and metabolism are increased in HF, likely due to FAO being reduced, and has also been proposed as treatment for HF^14,56,76,77^. We found no DA ketone bodies (3-hydroxybutyrate and acetoacetic acid) in the myocardium of end-stage HF patients. Instead, we found protein down-regulation of the intracellular monocarboxylate (ketone body) transporter 1 (SLC16A1), 3-hydroxybutyrate dehydrogenase (BDH1), and mitochondrial acetyl-CoA acetyltransferase (ACAT1) in ICM-DM, which are required for ketone metabolism to acetyl-CoA.

Proteins that catalyse BCAA metabolism to acetyl-CoA and succinyl-CoA, an alternative pathway to the citrate cycle, were also down-regulated in HF or exclusively in ICM-DM. This suggests down-regulation of BCAA metabolism in HF, particularly in ICM-DM, and may explain the up-regulation of myocardial BCAAs in HF, as previously indicated in end-stage NICM^14^, whereby increased tissue BCAA may impair insulin sensitivity^78^.

Myocardial Isoleucine leucine (all HF), leucine (ICM and NICM-DM), and valine (ICM) were up-regulated in ICM-DM. Increased plasma BCAA concentrations have been found in obese and type 2 DM patients, and DM-induced rodents with cardiomyopathy, and have been suggested to impair insulin-stimulated glucose uptake via the mTORC1 pathway^79–84^. Increased plasma and myocardial BCAAs have also been found in a diabetic cardiomyopathy mouse model with reduced expression of BCAT2, where pyridostigmine induced BCAA metabolism and clearance and attenuated the formation of myocardial scar tissue^85^.

Our finding of the down-regulation of OXPHOS subunits, particularly CI, in HF and has been reported previously in rodent and human HF and diabetes suggesting a more common HF pathway^86–91^. We found subunits of CI, CIII, CIV, and CV (Fig. 4a,e) with substantially greater significance and FCs in ICM-DM vs AMD relative to the other HF conditions. However, our modelling simulation incorporating CI changes alone demonstrated that reduced FCs are not a rate-limiting factor for sufficient NADH to NAD^+^ conversion/proton gradient, as suggested before^25–27^, but a limited capacity may be detrimental under stress^57,89^. Furthermore, down-regulated CI may result in other pathological consequences such as increased oxidative stress, hypertrophy, fibrosis, and glycolysis, and cell death via the mTOR signalling pathway (mTOR pathway proteins exclusively up-regulated in ICM-DM)^88,89,92,93^.

Evidence of this effect in our observations was seen in significant up-regulation of the NAD^+^/NADH ratio in HF paired with the down-regulation of NAD^+^ consumer sirtuin 3 (SIRT3)^94^. SIRT3 is a regulator of mitochondrial acetyl-CoA and OXPHOS via its deacetylation activity^89,95–102^. Our findings are in apparent contrast to previous studies reporting reduced NAD^+^/NADH in HF and in diabetes, mostly in hypertrophy HF rodent models, which may not translate to patients expressing end-stage HF prior to heart transplantation who do not have concentric cardiac hypertrophy but rather a dilated phenotype^89,103–105^. As high NAD^+^/NADH ratios have been coupled with tissue diacylglycerol accumulation via the NADH-dependent actions of glycerol-3-phosphate dehydrogenase, both of which are down-regulated in ICM-DM, our findings remain consistent^106,107^.

OXPHOS was further impaired in ICM-DM by the impairment of FADH_2_ oxidation in three pathways: (1) Oxidation via CII and the citrate cycle; (2) FAO FADH_2_ synthesis by the down-regulation of acyl-CoA dehydrogenases; and (3) Oxidation by GPD2 via glycerol metabolism^107,108^. This was supported in immunofluorescence microscopy by the apparent reduction of unbound flavins^32,34–37^. In addition, CoQ mediates the essential electron transfer from multiple sources (NADH, FADH_2_) between the biochemical actions of CI-CIII and is also a source of reactive oxygen species, while cytochrome C facilitates electron transfer to CIV^109^. We show CoQ subunits to be down-regulated specifically in ICM-DM, while COQ9 was down-regulated in DM.

Increased ROS formation from OXPHOS is principally due to pathological reverse electron transfer and it is associated with the metabolic dysfunction of DM^109,110^. The gene sets of iron-sulfur clusters, which mediate electron transfer in CI-CIII, were down-regulated specifically in DM^111^. Mitochondrial isocitrate dehydrogenase and malic enzyme 3 (ME3), which synthesise mitochondrial NADPH, were found to be down-regulated in ICM-DM. Though G6PD was up-regulated, it has been shown that there is no direct transport of NADPH from the cytosol to the mitochondria^112^. In concordance with our results, it has been found that reduction of CI results in reduced mitochondrial NADPH production and increased oxidative stress^93^.

Regarding intracellular ATP transport, we show down-regulation of the ATP/ADP antiporter, which transports ATP into the cytosol. We also found that kinases which increase ATP availability on demand, catalyse the formation of phosphocreatine, bind ATP, and act as an ATP energy buffer, were down-regulated in HF, particularly in ICM-DM, which is consistent with previous studies which also associated down-regulation of creatine kinase with a reduced OXPHOS capacity in HF, as well as overall reduced ATP availability^57,113^.

Mitochondrial dysfunction is not exclusive to diabetes and HF but also represented in other chronic diseases^114^. Yet, common protein markers used to predict mitochondrial volume/activity have been shown to be inadequate across different organs and in disease^28–31^. We propose SUCLG1, CKMT2, FH and ECHS1 as potential myocardial mitrochondrial protein biomarkers, as indicated by a reduced volume in ICM-DM which was immunobiologically verified by qualitatively reduced mitochondria.

Mitochondria are normally arranged in a regular, repeating pattern between myofibrils and between Z-discs and the cardiomyocyte cytoskeleton ^115–118^. We also observed disorganisation of mitochondria along the I-band, like the studies showing more numerous yet smaller mitochondria with cristae loss in ICM^90,119^ and diabetic^120^ rodent studies. However, we did not observe up-regulation in fission promoting proteins in HF in concordance with a human diabetes study which also reported no significant correlation between mitochondrial function and BMI^91,121^. To our knowledge, there have been no descriptions differentiating ICM with and without DM.

Considering the down-regulation of critical contractile complexes, we found that cardiac sarcoplasmic reticulum Ca^2+^ handling proteins ATPase and RYR2 were down-regulated in ICM-DM only. These have been identified to be reduced in HF previously, particularly the cardiac isoform of ATP2A2/SERCA2 wherein its affinity to Ca^2+^ and its expression were reduced^122–130^. In addition, Na^+^/Ca^2+^ exchanger SLC8A1 (Ca^2+^ efflux) was down-regulated specifically in ICM-DM, which contrasts with a study identifying increased left ventricular Na^+^/Ca^2+^ exchanger expression and activity in patients with end-stage HF^131^. SLC8A1 inhibitors have been proposed as a therapeutic target for HF as SAR340835 treated HF dogs showed an improved stroke volume and sympathovagal balance^132^. However, the positive effects in acute HF induced in animal models may not translate to chronic human end-stage HF^133^. We also identified the down-regulation of CASQ2 (Ca^2+^ binding) in DM, and the up-regulation of PLN, an inhibitory protein of ATP2A2. These changes of poor Ca^2+^ handling impede left ventricular relaxation and influence the rate of contraction in all end-stage HF conditions, but mostly in ICM-DM^127,134,135^.

Confocal microscopy revealed disorganisation of ATP2A2 along the I-band of the sarcomere in ICM-DM, co-localising with TNNI3. TNNI3 was down-regulated in ICM-DM and NICM suggesting further contractile impairment which is consistent with its release and elevation in the circulation of patients with advanced HF ^136–138^. Furthermore, it has recently been identified that nuclear translocation TNNI3 positively regulates ATP2A2 expression and Ca^2+^ uptake^139^.

ALDOA has been identified as a critical enzyme promoting glycolysis and as an F-actin binding protein at the I-band and a positive regulator of Ca^2+^ ryanodine receptors^38,140–142^. ALDOA is in low concentrations in post-MI patients’ plasma wherein an increased expression may have a protective role to alleviate oxidative stress and impair cardiomyocyte apoptosis in ischaemic conditions via the VEGF-NOTC1-JAG1 pathway^143^. However, overexpression of ALDOA has also been attributed to cardiac hypertrophy development^39^. Our study identifies that ALDOA co-localises at the I-band/Z-disc of human cardiomyocytes and is down-regulated and dislocated in ICM-DM, suggesting DM as a specific driver.

The expression of AEBP1, a protein known for its positive regulation of ECM organization and fibrosis-related proteins, including COL1A1 and COL1A2, which exhibited RNA-protein correlation/co-regulation in ICM-DM^144^, has been demonstrated to increase in response to hyperglycaemia, high fructose, and lipid concentrations in human hepatocytes^145^. The expression of AEBP1 has also been linked to diabetes^146^. AEBP1 has also been shown to act as a transcription factor that negatively regulates the gene expression of fatty acid binding protein, FABP4^147^. However, overexpression of AEBP1 in mice on a high-fat diet resulted in increased obesity^148,149^.

Myocardial up-regulation of the ECM and COL deposition via myofibroblasts are common maladaptive features in ICM and NICM^150–154^. We identified ICM-DM up-regulation of fibroblast activation protein alpha (FAP) transcript. Fibroblast integrin proteins, which promote ECM adhesion, can interact with POSTN, an ECM scaffolding protein, and stimulate angiogenesis, fibroblast proliferation and differentiation, and COL1 deposition by positively regulating TGFB1 and α-smooth muscle actin^155–159^. In addition, dermatopontin (DPT) positively regulates ECM formation including fibronectin and COL deposition and fibrillogenesis, which may promote maladaptive reverse remodelling in ICM and NICM^160–162^. Our study found DPT was up-regulated in all HF conditions with a greater than two-fold increase in ICM-DM relative to other HF conditions. This aligns with the previous finding of DPT and fibrosis up-regulation in adipocytes of individuals with obesity and obesity-associated type 2 DM^163^.

Taken together, we hypothesise a pathological feedback loop in the ICM-DM myocardium. ICM induced mitochondrial stress and an increase in ROS promoting lipogenesis may inhibit carnitine transport into the mitochondria and FAO, while also promoting the breakdown of lipid droplets in response to energy deprivation and the inhibition of adaptable glucose influx because of diabetes. The resulting intracellular lipid overload may contribute to impaired cardiac contraction, induced fibrosis, and an efflux of fatty acids from the cardiomyocytes. Further energy deprivation may be possible due to the apparent down-regulation of glycolysis, ketone and amino acid metabolism, and dysfunction of mitochondria.

This study is not without limitations. Efforts were made to control variables between conditions (age, macroscopic scar tissue, etc) to limit false results, including separate BMI and sex analyses where control was not possible. Acquisition and comparison of non-cardiomyopathy myocardium with diabetes was not possible. Not all proteins were quantified in this study, particularly low-abundance proteins, which may also have resulted in a lower RNA-protein correlation.

To our knowledge, this is the most comprehensive multi-omics resource on the molecular influence of DM in human end-stage ICM and NICM to date. Our results indicate that the cardiometabolic dysfunction of HF is further confounded and exacerbated in the presence of diabetes.

## Online Methods

### Acquisition of human myocardium

Pre-mortem human myocardium was acquired from the outer anterior and lateral walls of the left ventricle from explanted hearts of end-stage HF (ICM-DM, ICM-No DM, NICM-DM, and NICM-No DM) patients undergoing heart transplantation surgery, and from healthy/non-pathological donors whose hearts could not be viably used for heart transplantation due to logistical and compatibility limitations at the time. Procurement was performed at St Vincent’s Hospital Sydney as previously described^164–174^. Hearts were perfused in cardioplegia, placed on wet ice, and myocardial samples were immediately snap-frozen on-site in liquid nitrogen (-196°C) within 40 minutes after aortic cross-clamp such that the tissue was not post-mortem and that high-quality RNA was preserved^165,171,174^. Samples were then stored long-term at -192°C at the Sydney Heart Bank. Visible myofibrotic scar tissue, such as infarct-affected regions, were avoided. Pathology (HF) of myocardial tissue was histologically determined by hospital anatomical pathology such that non-pathological donor hearts did not present with disease. The methods of procurement, storage and use of donated human myocardium were approved by the Human Research Ethics Committee at The University of Sydney (USYD 2021/122). Due to limitations of local tissue availability at the time, all myocardial samples were Caucasian. Informed consent was obtained prior to the collection of all tissue and all ethical regulations relevant to human research participants were followed.

Dissemination of individual-level patient data is not permissible beyond condition/phenotype, age, sex, diabetes/no diabetes, and BMI. However, clinical and demographic data for each quantitative method has been provided in aggregate form (Supplementary Tables 1,43,55).

Clinical information was not available for every individual. Patient de-identified study IDs and the experiments each individual contributed to is available in Supplementary Table 59.

### Proteomics

Cryopreserved human myocardium fragments, lacking macroscopic myofibrotic scar tissue, were crushed and powederised with a mortar and pestle at -196°C, and weighed to ∼10mg. The powder was suspended in 4% sodium deoxycholate and 100mM Tris-HCl pH 7.5 and lysed in the ThermoMixerC (Eppendorf) at 95°C for 10 minutes. The homogenate was spun at 14,000 x g for 10 minutes and the supernatant collected. Stock protein concentration was quantified by performing a bicinchoninic acid (BCA, Pierce) assay and concentrations were corrected to 0.4 ng/µL. Protein was reduced in 10nM TCEP and 40nM chloroacetamide and denatured at 95°C for 10 minutes. A trypsin digest was then performed overnight at 37°C. Peptides were prepared for mass spectrometry as described previously^175^. A pooled sample was formed and was separated offline with high-pH RP fractionation. An acquity UPLC M-Class CSH C18 130 Å pore size, 300 µm x 150 mm column, and 1.7 µm internal diameter (Waters) was used to fractionate peptides. Separation was performed over 30 minutes, concatenating 96 fractions into 16 wells using buffer A; 2% acetonitrile and 10 mM ammonium formate (pH 9), and buffer B; 80% acetonitrile and 10 mM ammonium formate (pH 9). Fractionates were dried and resuspended in 5% formic acid. Peptides were directly injected into a C18 (Dr. Maisch, Ammerbuch, Germany, 1.9 µm) 30cm x 70cm column with a 10 µm pulled tip integrated online to a nanospray ESI source. Separation involved buffer A; 0.1% formic acid in liquid chromatography (LC)-MS grade water, and buffer B; 80% acetonitrile and 0.1% formic acid in MS grade water. Peptides were differentiated from a gradient of 5% -40% of buffer B over 2 hours with a flow rate of 300 nL per minute and were ionised by electrospray ionisation at 2.3 kV. A Q-Exactive Fusion Lumos mass spectrometer (ThermoFisher) was used to conduct MS/MS analysis with 27% normalised HCD collision energy for fragmentation. Spectra were procured in a data-independent acquisition using 20 variable isolation windows. RAW data files including the high pH fractions were analysed using the integrated quantitative proteomics software DIA-NN^176^ (version 1.7). The database provided to the search engine for identification contained the Uniprot human database downloaded on the 5th of May, 2020. FDR was set to 1% of precursor ions. Both remove likely interferences and match between runs were enabled. Trypsin was set as the digestion enzyme with a maximum of 2 missed cleavages. Carbamidomethylation of Cys was set as a fixed modification and oxidation of Met was set as variable modifications. Retention time-dependent profiling was used, and the quantification setting was set to any LC (high accuracy). Protein inference was based on genes. The neutral network classifier was set to double-pass mode. The MaxLFQ algorithm was used for label-free quantitation, integrated into the DIA-NN environment^177,178^.

Normalised and log_2_ transformed proteomic data per individual is available in Supplementary Table 60.

### Metabolomics

Cryopreserved human myocardium was powderised as described in the proteomic MS methods and weighed to 50mg. Cells were lysed and homogenised with steel balls in methanol/chloroform (2:1; v/v, HPLC grade) using the TissueLyser LT (Qiagen) and kept on dry ice between cycles (3-5 x 1 minute). Equal volumes of chloroform then water (HPLC grade) were added to promote protein precipitation. Protein and debris were pelleted at 14,000 rpm at 4°C for 20 minutes and the metabolite-containing supernatant was collected.

The Speed-Vac SPD120 (Thermo Fisher Scientific) was used to dry the solution under nitrogen steam. Compounds were resuspended in the acetonitrile/methanol/formic acid (75:25:0.2; v/v/v, HPLC grade; Thermo Fisher Scientific) for the HILIC analysis, and acetonitrile/methanol (25:25; v/v/v, HPLC grade) for the AMIDE analysis. Targeted metabolite profiling was performed using deuterated internal standards to determine mass spectrometry (MS) multiple reaction-monitoring transitions, declustering potentials, collision energies and chromatographic retention time, as described previously^179,180^. HILIC and AMIDE mass spectrometry analyses were performed via liquid chromatography-tandem mass spectrometry (LC-MS/MS) using an Agilent 1260 Infinity liquid chromatography (Santa Clara, CA, USA) system coupled to a QTRAP5500 mass spectrometer (AB Sciex, Foster City, CA, USA). Positive and negative ion modes were used to separate polar compounds in hydrophilic interaction liquid chromatography (HILIC) mode using an Atlantis® HILIC column (Waters), and an XBridgeTM Amide column (Waters), respectively, which allow the separation of metabolites of different properties, as previously described^179,180^. Samples were randomised across three sequential batches. Pooled metabolite extracts formed from each sample were included after every 10 study samples in the sample queue to detect temporal dips in instrument performance following the analysis. SCIEX OS (AB SCIEX, version 1.7.0.36606) was later used for multiple reaction monitoring Q1/Q3 peak integration of the raw data files whereby the abundance was quantified as the peak area of a metabolite. QC was performed removing metabolites with poor quantification or to select between metabolites measured in both negative and positive modes. Metabolites abundances were normalised in R (version 4.3.1.) using scMerge (version 1.16.0)^181^.

Normalised and log_2_ transformed metabolomic data per individual is available in Supplementary Table 61.

### Lipidomics

Cryopreserved human myocardium was powderised as described in the proteomic MS methods and weighed to 20mg. Internal standards were added to each sample in 250 µL methanol (MS grade) containing 0.01% (w/v) butylhydroxytoluene (BHT); 2 nmoles PC(19:0/19:0); 1 nmole each of SM(d18:1/12:0), GluCer(d18:1/12:0), Cer(d18:1/17:0), PS(17:0/17:0), PE(17:0/17:0), PA(17:0/17:0), PI(d7-18:1/15:0), PG(17:0/17:0), CL(14:0/14:0/14:0/14:0), and TG(17:0/17:0/17:0); 0.5 nmoles each of DG(d7-18:1/15:0), CholE(17:0), LPC (17:0), LPE(17:1), and AcCa(d3-16:0); and 0.2 nmole each of Sph(d17:1), S1P(d17:1), LacCer(d18:1/12:0), and MG(d7-18:1). Samples were then homogenised using steel balls in the TissueLyser LT (Qiagen) and kept on wet ice between cycles (3-5 x 1 minute). Methyl-tert-butyl-ether (MTBE, 1 mL) was added to the homogenate and samples sonicated at 4°C for 30 minutes. Phase separation was induced by adding 250 µL of water (MS grade), vortexing and spinning at 2,000 x g for 5 minutes. The upper organic phase was extracted in 5 mL glass tubes. Extraction was performed twice more; sonicated at 4°C for 30 minutes in 500 µL MTBE and 150 µL methanol, then vortexed and spun at 2,000 x g for 5 minutes after adding 150 µL water. Lipids were dried under vacuum in a Savant SC210 SpeedVac (ThermoFisher Scientific) and reconstituted in 400 µL 80% methanol/20% water/0.1% formic acid containing 0.01% (w/v) BHT. Lipids were then analysed via liquid chromatography-tandem mass spectrometry (LC-MS/MS). LC-MS/MS was performed using a Q-Exactive HF-X mass spectrometer (ThermoFisher) coupled to a Vanquish HPLC with a 2.1x100 mm C18 HPLC column (Waters, 1.7 µm pore size) as previously described^182^ with amendments in the following. HPLC solvent A was 10 mM ammonium formate, 0.1% formic acid in acetonitrile:water (60:40), and solvent B was 10 mM ammonium formate, 0.1% formic acid in isopropanol:acetonitrile (90:10). A 27 min binary gradient at 0.28 mL/min was used: 0 min, 80:20 A/B; 3 min, 80:20 A/B; 5.5 min, 55:45 A/B; 8 min, 36:65 A/B; 13 min, 15:85 A/B; 14 min, 0:100 A/B; 20 min, 0:100 A/B; 20.2 min, 70:30 A/B; 27 min, 70:30 A/B.

Data was acquired in full scan/data-dependent MS2 mode (full scan resolution 60,000 FWHM, scan range 220–1600 m/z). Sample order was randomised, and data was collected in both positive and negative mode for each sample. The ten most abundant ions in each cycle were subjected to MS2, with an isolation window of 1.4 m/z, collision energy 30 eV, resolution 17,500 FWHM, maximum integration time 110 ms and dynamic exclusion window 10 s. A solvent blank was used to created an exclusion list of background ions while an inclusion list was used for all internal standards. LipidSearch software (version 4.2, Thermo Fisher) was used for lipid annotation, chromatogram alignment, and peak integration. Lipid annotation required both accurate precursor ion mass (tolerance 5 ppm) and diagnostic product ions (tolerance 8 ppm). Individual lipids were expressed as ratios to the class-specific internal standard, then multiplied by the amount of internal standard to calculate molar amounts for each lipid. Lipid levels were expressed as nmoles/mg tissue.

Normalised and log_2_ transformed metabolomic data per individual is available in Supplementary Table 62. Lipid class names are available in Supplementary Table 63.

### RNA sequencing

Cryopreserved human myocardium was powderised as described in the proteomic MS methods and weighed to approximately 30mg. Powderised myocardium was lysed and homogenised in 500 µL TRIzol (Invitrogen) using steel balls in the TissueLyser LT (Qiagen) and kept on dry ice between cycles (3-5 x 1 minute). 1-bromo-3-chloropopane (50 µL) was added to each sample and left to incubate for 5 minutes at RT, then spun at 14,000 x g at 4°C for 15 minutes to induce phase separation. RNA-containing aqueous phase was transferred to a sterile tube and an equal volume of isopropanol was added, mixed by inverting, and left for 1 hour at RT. RNA was pelleted at 14,000 x g at 18°C for 15 minutes, supernatant discarded. RNA was then washed in 4°C 70% ethanol twice discarding supernatant after spinning at 14,000 x g at 4°C for 10 minutes and left to dry at RT. RNA was DNase treated for 30 minutes at 37°C and washed again. Pelleted RNA was reconstituted in 15 µL 4°C nuclease free diethyl pyrocarbonate (DEPC). Concentration and quality were tested on NanoDrop (Thermo Fisher Scientific) to a standard of 260nm/280nm = 1.8-2.0 and 260nm/230nm 1.7-2.2. RNA integrity was assessed using an RNA Nano Chip on an Bioanalyzer (Agilent).

RNA-seq libraries were prepared with Illumina Stranded Total RNA prep Ligation with Ribo Zero Plus (100ng input and 11 PCR cycles) according to manufacturer’s instructions. Quality checks were performed using the Qubit dsDNA High Sensitivity Assay Kit and the Revvity LabChip GX Touch. The final sequencing pool was quantified using the Qubit dsDNA High Sensitivity Assay Kit after pooling all libraries equimolar into a single library pool. Sizing was checked using the Agilent High Sensitivity D1000 Tapestation system. The RNA-seq libraries were sequenced using a paired-end 250bp kit on a S4 flow cell of the Illumina NovaSeq 6000 with a final run concentration of 58pM and 1% PhiX. The raw data was demultiplexed using bcl2fastq. Library preparation and sequencing were performed by the Ramaciotti Centre for Genomics, at the University of New South Wales, Australia. The quality of each RNA sequence (reversely stranded) was assessed using ShortRead and the alignment was performed on paired-end sequences without trimming^183,184^. The alignment of raw sequences was performed on hg38 genome assembly (UCSC) with Rsubread version 2.2.1^184,185^. Rsubread was also used for generating the gene count matrix. The gene counts were converted in Count Per Million (CPM), log_2_ transformed, and then normalised using voom^186^.

Normalised and log_2_ transformed RNA sequence data per individual is available in Supplementary Table 64.

### Statistics and reproducibility

Differential abundance (DA) or differential expression analyses were performed on proteomic, metabolomic, lipidomic and RNA-seq data sets using a moderated t-test with the limma package (version 3.56.2) in R (version 4.3.1) following log_2_ transformation. HF conditions (ICM and NICM) were compared with AMDs; Donors aged 47-65 years for ICM, and Donors aged 21-65 years for NICM. DA analyses were also performed for ICM-DM was also compared against ICM-No DM, and males against females. Molecules with missing values in more than 25% of samples were excluded. No imputation method was applied. Significant proteins, metabolites, and lipids were determined by controlling for False Discovery Rate at the 5% level using Benjamini-Hochberg FDR adjustment (FDR < 0.05) and ranked using the topTable function in limma. In the RNA-seq differential analysis, transcript significance was defined as unadjusted significant (*P* = 0.05) and > ±2-FC. Differential analyses are located in Supplementary Tables 2-20, 44-46, 52, 56-58.

Correlation between RNA and protein (RNA-protein), BMI and molecule, and individual mitochondrial protein expression and the average MitoCarta3.0 protein expression were calculated using Pearson’s correlation coefficient whereas the correlation between male and AEBP1 fold change (FC, HF compared to AMD) was calculated using Spearman’s correlation coefficient. Correlation between individual mitochondrial protein expression and the total average expression of the mitochondrial proteome (MitoCarta3.0) were also calculated via Pearson’s correlation coefficient where the mitochondrial proteome was limited to proteins quantified and proteins expressed by all samples. As protein expression was determined via relative quantification, skewing of disproportionally abundant proteins was dampened, however, expression of highly abundant proteins would be resistant to pathological changes *in vivo*. Correlation was determined as ρ > 0.8 (Pearson, exceeded FDR), or as ρ > 0.8 and *P* < 0.05 (Spearman) in the male/sex influenced analysis (Supplementary Fig. 9g). Correlation analyses are located in Supplementary Tables 37-42, 48-51.

Gene Set Enrichment Analysis (GSEA) analyses were performed on normalised and transformed proteomic and metabolomic datasets using clusterProfiler (version 4.8.1)^187^ in R where significance was determined as *P* < 0.05 (unadjusted). Protein enrichment was performed referencing the databases; Kyoto Encyclopedia of Genes and Genomes (KEGG, accessed 21^st^ June 2022)^188–190^ via KEGGREST (version 1.40.0), MitoCarta3.0^191^, and Gene Ontology (GO, version 3.17.0) Biological process (BP) and Cellular Component (CC)^192,193^. Selected mitochondrial and energetics-associated KEGG pathways and MitoCarta3.0 gene sets were manually annotated. The Kolmogorov-Smirnov statistic was used to compare the ranks of *P* values of genes in the pathways/gene sets vs the uniform distribution using the “GSEA” function. GSEA GO enrichment results were the child nodes of manually selected cytoskeletal, sarcomeric, and extracellular matrix-associated parent nodes; extracellular matrix organisation (GO:0030198), cardiac muscle cell differentiation (GO:0055007), myofibril assembly (GO:0030239), heart contraction (GO:0060047), cytoskeleton organization (GO:0007010), extracellular matrix (GO:0031012), contractile fibre (GO:0043292), and cytoskeleton (GO:0005856). Significantly enriched protein pathways/gene sets/nodes were fully represented (NICM) or manually selected (ICM) in bubble plot figures. Metabolite enrichment was performed referencing on the KEGG database. clusterProfiler enrichment analyses are located in Supplementary Tables 21-32.

Enrichment analyses of targeted genes were performed following and based on the FDR accepted proteomic and RNA-seq differential abundance analyses results using GO via PANTHER (version 17.0)^194^ and STRING (version 12.0)^195,196^ where significance was determined as FDR < 0.05. STRING analysis (version 11.5) was also performed on ICM-DM RNA-protein correlated genes. GO via PANTHER analyses referenced BP, CC, and Molecular Function (MF) libraries where inputted data was direction specific (down or up-regulated) for each analysis before combining the results. STRING analyses referenced GO (BP, CC, and MF) and KEGG databases. Significantly enriched nodes/pathways were manually selected for the figures. Targeted enrichment analyses are located in Supplementary Tables 33-36, 47, 53, 54.

Figure elements were generated in R using ggplot2 (version 3.4.2), clusterProfiler, GraphPad Prism (version 9.5.1), DeepVenn^197^, Microsoft Excel (version 2309), Adobe Illustrator (version 27.8), at https://app.biorender.com, and https://www.bioinformatics.com.cn/en.

Molecules of the circular heat maps were clustered via complete-linkage hierarchical clustering, and Log_2_ FCs were separated via Euclidean distance. RNA-protein scatter plots only plotted genes with a relative RNA expression ≥ 5. CC nodes of the GO chord plot were manually selected based on enrichment. UpSet plots were produced using FDR significant proteins/metabolites in HF vs AMD from the differential analyses. Elements were modified and formatted using Adobe Illustrator.

### Verhoeff-Van Gieson histochemistry & bright-field microscopy

Verhoeff-Van Gieson staining was used to primarily stain collagen and elastin fibres. Donor and ICM-DM cryopreserved pre-mortem human left ventricular myocardium was sectioned on the Cryotome FSE (Thermo Fisher Scientific) at -16°C while in tissue freezing medium to 16 µm thickness and collected onto Superfrost Plus slides (Menzel, Thermo Fisher Scientific). Sections were fixed in methanol for 3 minutes at -20°C, immersed in tap water. Elastin fibres and nuclei were stained in Verhoeff’s Iron Haematoxylin solution; 2.8% filtered ethanol haematoxylin (w/v), 22% ferric chloride (v/v), 22% strong iodine, for 25 minutes then differentiated in 2% aqueous ferric chloride for 10 dips/seconds. Iodine was washed out in 95% ethanol for 30 seconds and collagen was then counterstained in Van Gieson’s stain; 0.1% aqueous acid fuchsin, in 90% saturated picric acid (v/v), for 3 minutes. Sections were then dehydrated in ascending changes of ethanol, cleared in xylene, and mounted in DPX (Sigma-Aldrich).

Bright-field images (24 bid depth) were captured on ZEN blue (version 3.3) using the Zeiss Axio Scan.Z1 and a 40x plan-apochromat objective with a numerical aperture (NA) of 0.95 on the Hitachi HV-F202SCL camera with an exposure time of 200 µs. Images were adjusted (brightness and contrast) to identical ranges and exported in Tag Image File Format (TIFF) using Fiji/ImageJ^198^.

### Immunohistochemistry & confocal microscopy

Donor and ICM-DM sections were sectioned as described for Verhoeff-Van Gieson staining. Sections were encircled with a PAP pen and fixed in 10% neutral buffered formalin for 15 minutes then washed from free aldehyde groups in 0.2% glycine (w/v) in phosphate buffered saline (PBS). Slides were washed for 5 minutes three times in PBS between steps at 40 RPM. Tissue was permeabilised in 0.5% (v/v) Triton X-100 in PBS before non-specific binding sites were blocked in 5% (v/v) normal donkey serum and 5% acetylated bovine serum in PBS for 45 minutes at room temperature (RT) at 40RPM. Monoclonal primary antibodies (IgG) produced in rabbit labelling TNNI3 (ab47003, ∼5 µg/mL, abcam), NDUFS3 (ab177471, ∼20 µg/mL, abcam), GFM1 (HPA061405, ∼16 µg/mL, Sigma-Aldrich), or ALDOA (HPA004177, ∼10 µg/mL, Sigma-Aldrich) were applied to the sections in 10% blocking solution overnight at 4°C at 40 RPM. Rabbit IgG were in-turn labelled with secondary fluorophore-conjugated Alexa Flour 488 IgG (ab15065, 5 µg/mL, abcam) overnight at 4°C at 40 RPM. Co-labelling was similarly performed in sequence with mouse-produced monoclonal IgG labelling ATP2A2 (MA3919, ∼10 µg/mL, Invitrogen) or ACTN2 (A7732, ∼20 µg/mL, Sigma-Aldrich) and secondary IgG DyLight 550 (ab9879, 5 µg/mL, abcam). Plasma/intra-membrane glycoproteins were bound with Alexa Flour 647-conjugated wheatgerm agglutinin (W32466, 5 µg/mL, Molecular Probes) for 1 hour at RT at 40 RPM. Nuclei were stained with DAPI (62248, 1 µg/mL, Thermo Fisher Scientific) for 10 minutes at RT at 40 RPM. Sections were then mounted in ProLong Diamond antifade mountant (Thermo Fisher Scientific).

Z-stack images (12 bit depth) with a thickness of 4.5 µm and step size of 0.3 µm (16 images) were captured on NIS-elements (version 4.0) using a Nikon C2plus confocal microscope, a 100x oil-immersed plan-apochromat objective with a NA of 1.45 and image parameters; 1.0 airy unit (AU), 1.3 x digital zoom, 1/16 frames/sec, 2 x frame-average, and a resolution of 2048 x 2048. Channels were imaged in sequence from long to short excitatory laser wavelengths; 640 nm (emission filter 650 – 700 nm), 561 nm (emission filter 570 – 620 nm), 488 nm (emission filter 500 – 550 nm), and 405 nm (emission filter 417 – 477 nm).

Acquisition parameters including laser power, gain, and offset were identical between donor and ICM-DM and no-primary IgG controls (ICM-DM). All channels were deconvolved with Huygens Profesional version 22.10.0p1 64b (Scientific Volume Imaging, The Netherlands, http://svi.nl), using the CMLE algorithm. Stain-specific templates were used between conditions to ensure identical deconvolution parameters (acuity, signal to noise ratio, iterations, and background value). Z-stacks were compressed into a two-dimensional image using Maximum Intensity Projections, adjusted (brightness and contrast) to identical ranges and exported in OME-TIFF using Fiji/ImageJ^198^.

### In-silico modelling

To study the physiological consequences of reduced complex I (CI) activity, we employed a biophysical model of oxidative phosphorylation validated on rat cardiac myocytes^199–201^. Equations of the model were sourced from Malecki et al (2023, In-Review). We reduced the activity parameter corresponding complex I (*μ*^*α*^_C1_) according to the equation

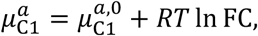

where *μ*^*α*,0^_C1_ = -21.01 kJ/mol is the baseline value of the activity parameter, *R* = 8.314 J/K/mol is the ideal gas constant, *T* = 297.5 K is the absolute temperature, and FC is the fold change in CI abundance relative to the baseline abundance. The parameters corresponding to the groups were:

- AMD: Baseline value (FC = 1.0).
- ICM-DM: -29.4% from baseline (FC = 0.706), consistent with fold changes in the proteomic data. The fold change was calculated from the mean of the fold changes (ICM-DM relative to AMD) in the CI subunits present in the proteomic data. CI subunits were defined using the HUGO Gene Nomenclature Committee (HGNC) groups

“NADH:ubiquinone oxidoreductase core subunits” (https://www.genenames.org/data/genegroup/#!/group/1149) and “NADH:ubiquinone oxidoreductase supernumerary subunits” (https://www.genenames.org/data/genegroup/#!/group/1150).

- Low C1: -40% from baseline (FC = 0.6). This group was included to study the effects of reducing complex I beyond the fold changes seen in the proteomic data.

The model was simulated for each group by iteratively varying the ATP consumption parameter *μ*^*α*^_ATPase_, leading to changes in the oxygen consumption rate VO_2_ = 0.5*J*_C4_. For each value of *μ*^*α*^_ATPase_, the model was simulated for 1000 seconds to reach steady state.

## Data availability

Proteomic mass spectrometry raw data is available at ProteomeXchange via PRIDE with the project accession PXD052878. Reviewers should use the token, ‘NhmhQafIt5jS’. Reviewers can also access using the username, ‘’, with the password, ‘gEWhZ1N6VXPO’.

Metabolomic mass spectrometry raw data is temporarily available at Metabolomics Workbench with the Study ID ST003274 and Project DOI PR002031 at https://dev.metabolomicsworkbench.org:22222/data/DRCCMetadata.php?Mode=Study&StudyID=ST003274&Access=DleC4637. When released, the data with be available at https://dx.doi.org/10.21228/M83529.

Lipidomic mass spectrometry raw data is temporarily available at Metabolomics Workbench with the Study ID ST003275 and Project DOI PR002031 at https://dev.metabolomicsworkbench.org:22222/data/DRCCMetadata.php?Mode=Study&StudyID=ST003274&Access=DleC4637. When released, the data with be available at https://dx.doi.org/10.21228/M83529.

RNA sequencing raw data is available at NCBI Gene Expression Omnibus (GEO) at https://www.ncbi.nlm.nih.gov/geo/query/acc.cgi?acc=GSE263297. Reviewers should use the token, ‘cvsfeoeevxglloh’.

## Code availability

Code is available at https://zenodo.org/records/14048213. DOI: 10.5281/zenodo.14048213

## Author contribution

B.H. designed the study and the sample cohort, performed sample preparation and extraction for proteomic, metabolomic, lipidomic, and RNA-seq analyses, performed metabolite quantification, assisted with differential analyses, co-designed and performed statistical analyses, performed histochemistry, brightfield and confocal microscopy, analysed the differential and statistical results, performed biological interpretation, produced the figures and figure legends, and wrote the manuscript. Y.Z. Normalised the metabolomic MS data, performed differential analyses, co-designed and performed statistical analyses, assisted with designing, and producing the figures, and co-developed the shiny app. D.H. performed proteomic MS, protein quantification and normalisation. H.M. performed lipidomic MS, lipid quantification and normalisation. Y.C.K. performed metabolomic MS. M.P. performed oxidative phosphorylation computational model simulations. C.M. assisted with RNA extraction. J.K. performed lipid quantification and normalisation. R.H. supervised confocal microscopy. G.G. processed RNA-seq raw data. A.W. co-developed the shiny app. S.D. co-developed the shiny app. V.R. supervised M.P. A.D. supervised H.M. and J.K. M.L. supervised D.H. J.F.O.S. supervised Y.C.K. and edited the manuscript. J.Y. lead the bio-statistical analyses and supervised Y.Z. in the above. S.L. conceived and led the study, edited and contributed to the manuscript, co-designed the figures, and supervised B.H. in the above.

## Acknowledgements

We thank the patients and staff of St Vincent’s Hospital, Sydney, and the Australian Red Cross Blood Service. We thank Emeritus Prof. Cris dos Remedios and the late Dr. Victor Chang AC who established the Sydney Heart Bank. We thank Ben Crossett and Stuart Cordwell from Sydney Mass Spectrometry. This work was funded by the R.T. Hall Trust (RQ112) and philanthropic donations to the University of Sydney. This work was also supported by the National Health and Medical Research Council (NHMRC) of Australia and the National Heart Foundation (NHF) of Australia. The contents of the published material are solely the responsibility of the individual authors and do not reflect the view of NHMRC or the NHF. J.O.S. is supported by an NSW OHMR EMCR grant (G212785), NHF Vanguard Grant (105595), NHF Future Leader Fellowship (104853), and an NHMRC/MRFF CVD award (2024161). M.P. was supported by a Postdoctoral Research Fellowship from the School of Mathematics and Statistics, University of Melbourne. Metabolomics Workbench raw data availability is supported by NIH grant U2C-DK119886 and OT2-OD030544 grants.

## Competing interests

The authors declare no competing financial or non-financial interests.

---

**Supplementary Figure 1.**
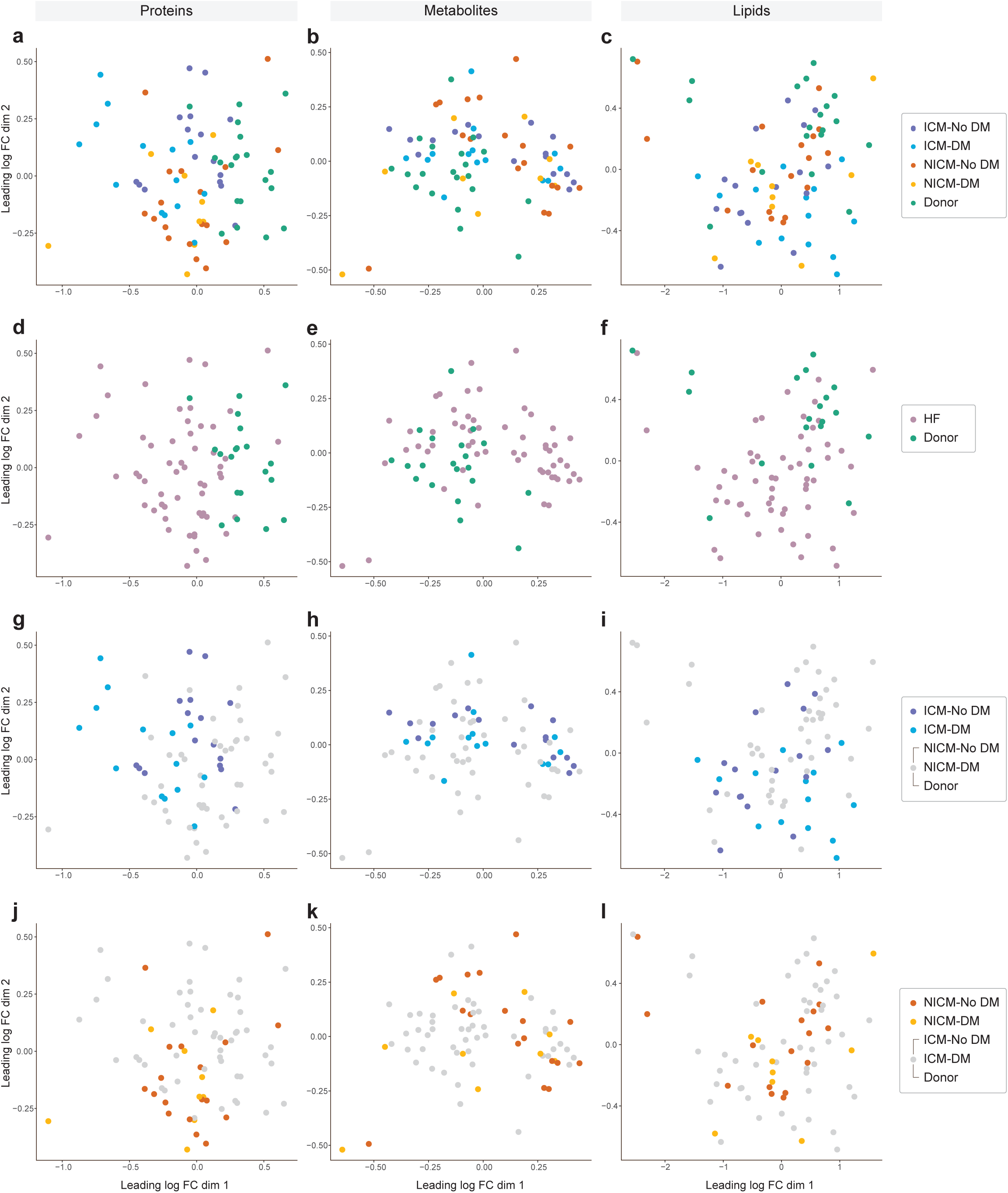
Sample visualisation of normalised and log_2_ transformed proteomic, metabolomic and lipidomic mass spectrometry data from human myocardium in all conditions. **a**-**l**, Multidimensional scaling (MDS) plots, a tool to visualise high dimensional data in two dimensions, were used to iteratively separate samples (each point) in distance based on their dissimilarity of paired data (proteins, metabolites, and lipids; dimensions) calculated as the leading log_2_ fold change (FC, average root-mean-square of the largest log_2_ FCs). All protein/metabolite/lipid MDS plots are the same but vary in samples highlighted. **a**-**c**, All conditions highlighted. **d**-**f**, Heart failure (HF) conditions and healthy donors highlighted. **g**-**i**, Ischaemic cardiomyopathy with diabetes (ICM-DM) and without diabetes (ICM-No DM) highlighted. **j**-**l**, Non-ischaemic cardiomyopathy with diabetes (NICM-DM) and without diabetes (NICM-No DM) highlighted.

**Supplementary Figure 2.**
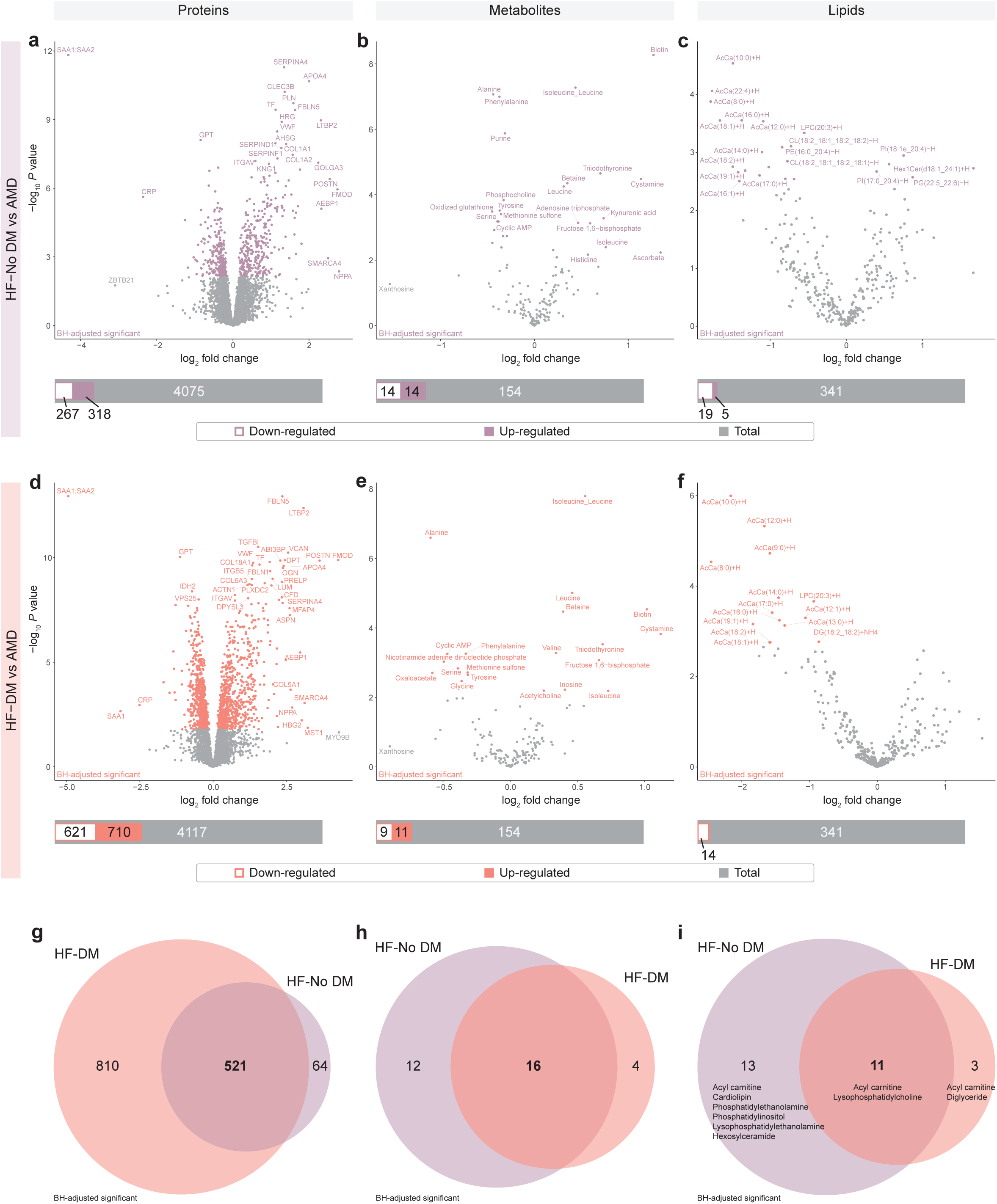
Human left ventricular myocardial differential analysis in protein, metabolite, and lipid abundance between heart failure with (HF-DM) and without diabetes (HF-No DM) and age-matched donors (AMD). **a-f**, Differential abundance was determined following Benjamini-Hochberg false discovery rate adjustment (FDR) of *P* values (FDR < 0.05). Superimposed bar plots summarise the number of significantly down-regulated (white-filled bar) and up-regulated (colour-filled bar) molecules relative to the total number of molecules analysed (grey bar). Down and up-regulation was defined as significance of molecule abundance in the heart failure condition compared to healthy age-matched donors (AMD). **a**-**c**, HF-No DM vs AMD. **d**-**f**, HF-DM vs AMD. **g**-**i**, HF-DM and HF-No DM vs AMD FDR significant differentially abundant proteins, metabolites, and lipids, respectively. **i**, Lipid classes annotated in order of the number of lipids which were FDR significant in that particular class.

**Supplementary Figure 3.**
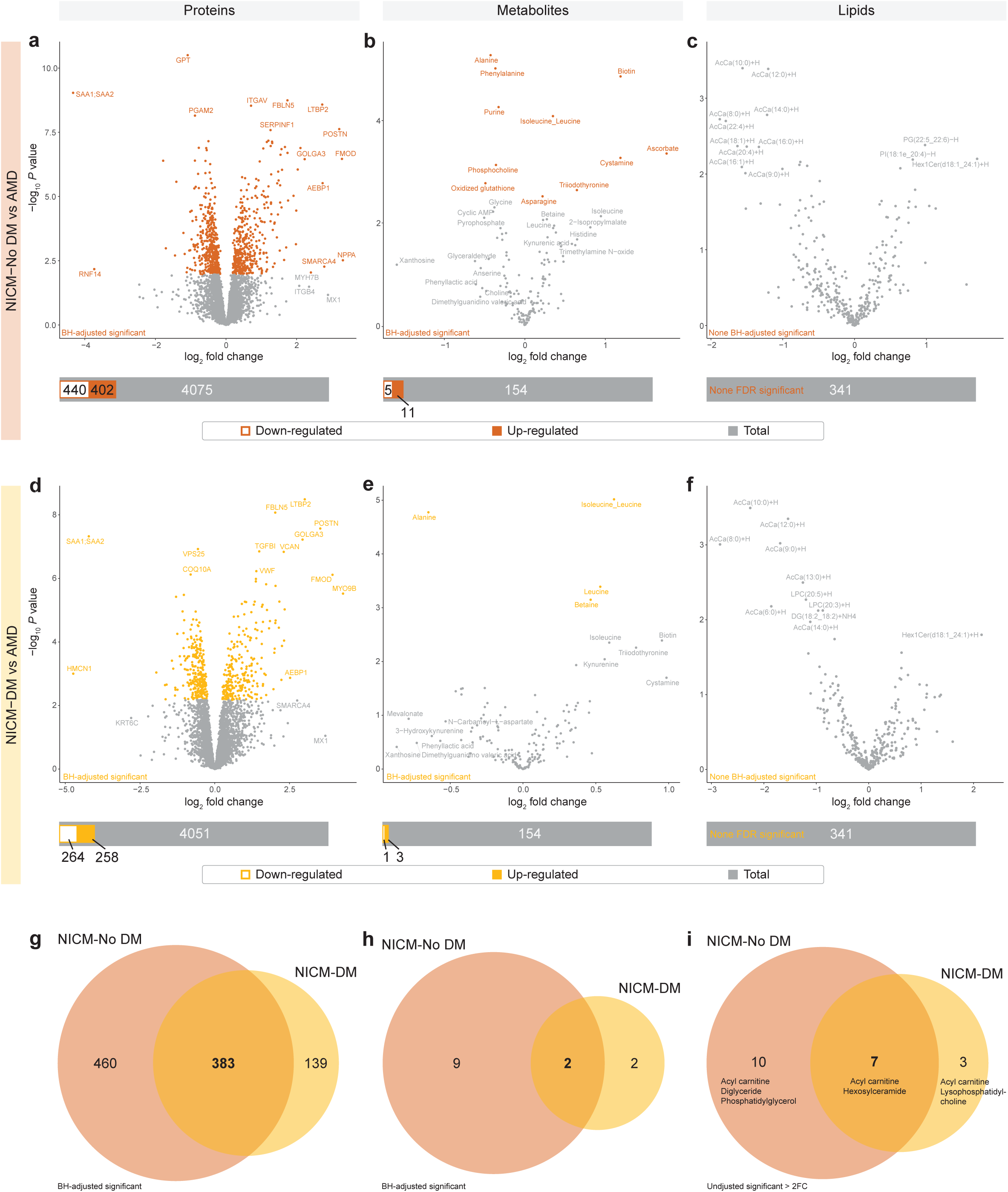
Human left ventricular myocardial differential analysis in protein, metabolite, and lipid abundance between non-ischaemic cardiomyopathy with (NICM-DM) and without diabetes (NICM-No DM) and age-matched donors (AMD). **a-f**, Differential abundance was determined following Benjamini-Hochberg false discovery rate adjustment (FDR) of *P* values (FDR < 0.05). Superimposed bar plots summarise the number of significantly down-regulated (white-filled bar) and up-regulated (colour-filled bar) molecules relative to the total number of molecules analysed (grey bar). **a**-**c**, NICM-No DM vs AMD. **d**-**f**, NICM-DM vs AMD. **g**-**h**, NICM-DM and NICM-No DM vs AMD FDR significant differentially abundant proteins and metabolites. **i**, NICM-DM vs AMD and NICM-No DM vs AMD unadjusted significant (*P <* 0.05) lipids which were also > ±2-FC (in either direction) for comparative purposes (significance not accepted). Lipid classes annotated in order of the number of lipids which were unadjusted significant.

**Supplementary Figure 4.**
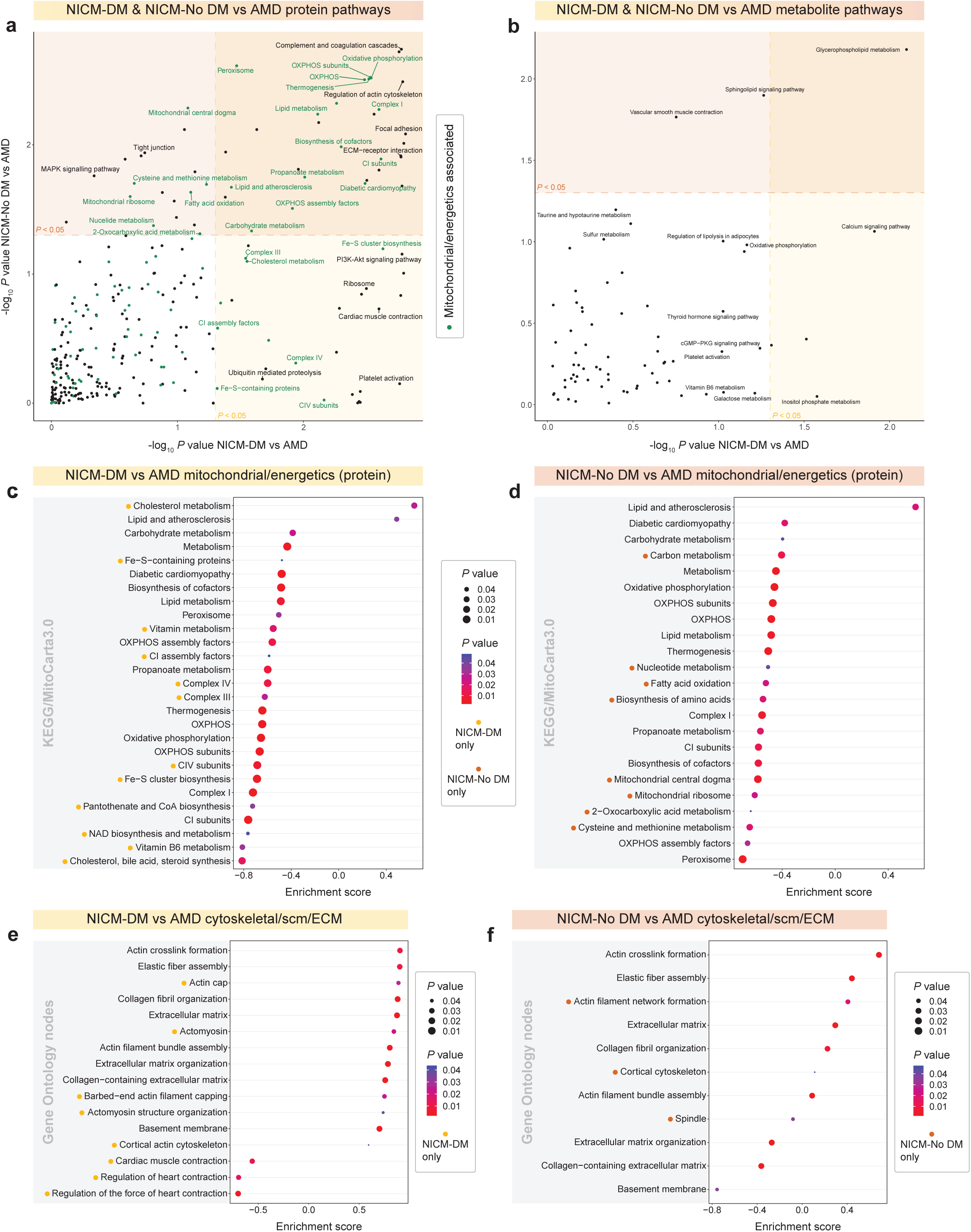
Human non-ischaemic cardiomyopathy with (NICM-DM) and without diabetes (NICM-No DM) vs age-matched donor (AMD) myocardial protein and metabolite pathway analyses. **a**-**b**, Scatter plots of -log_10_ *P* value enriched KEGG and MitoCarta3.0 protein pathways/gene sets of NICM-DM AMD and NICM-No DM where mitochondrial and energetics related pathways are coloured in green and significant (*P* < 0.05). Pathways in overlapping coloured regions were significant in both NICM-DM and NICM-No DM vs AMD. **a**, Enriched pathways from the proteomic mass spectrometry (MS) analysis. **b**, Enriched pathways from the metabolomic MS analysis. **c**-**f**, Gene Set Enrichment Analysis (GSEA) enrichment bubble plots of top ranked significantly enriched (*P* < 0.05) KEGG/MitoCarta3.0 pathways/gene sets and Gene Ontology nodes, respectively. **c**-**d**, From Fig. S4a. **e**-**f**, Cytoskeletal, sarcomeric (scm), and extracellular matrix (ECM) enriched Gene Ontology Biological Process and Cellular Component nodes from selected parent nodes.

**Supplementary Figure 5.**
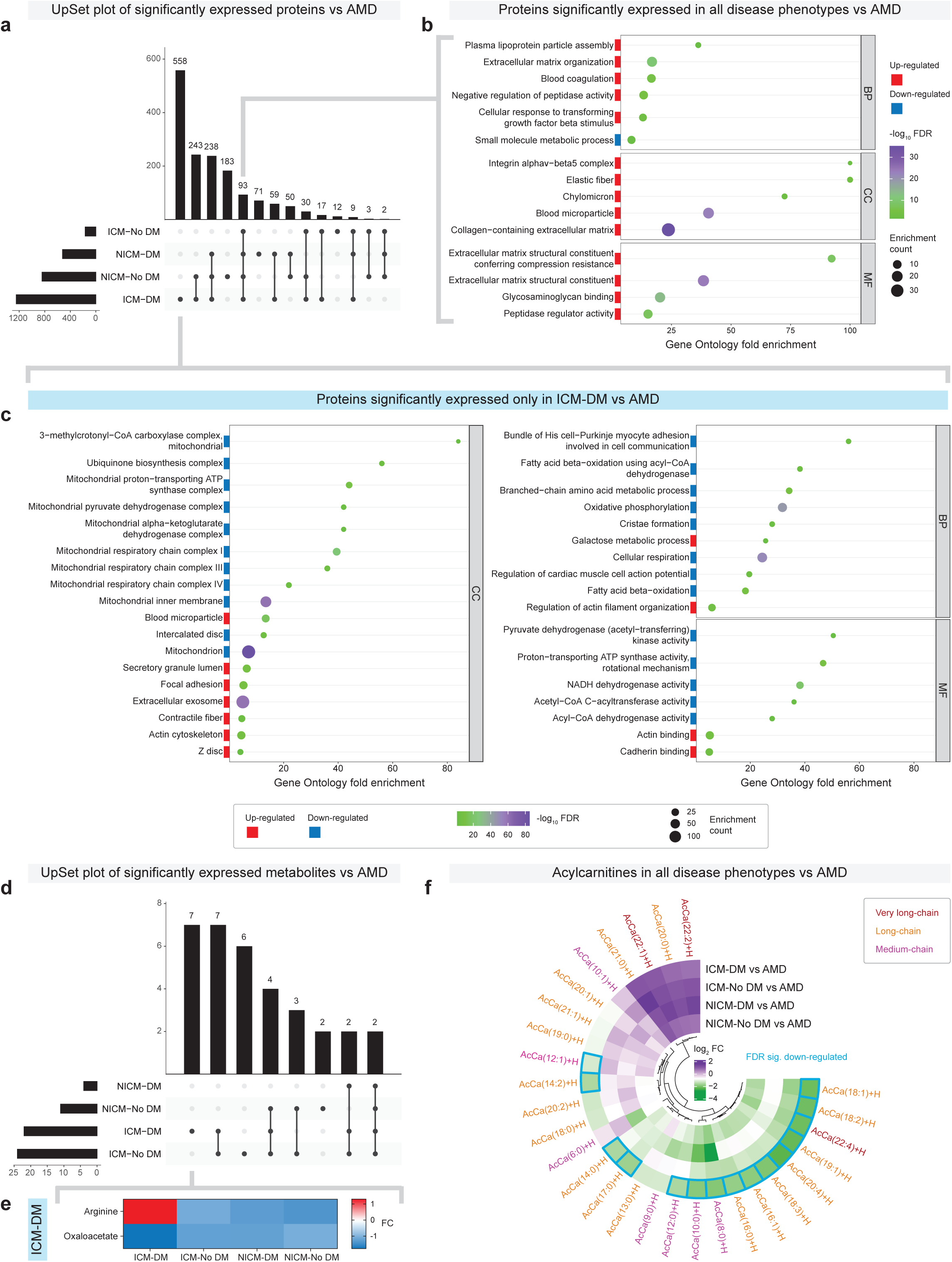
Myocardial differential abundance co-analyses of all heart failure conditions vs age-matched donors (AMD), relative to each other, for proteomic, metabolomic, and lipidomic mass spectrometry analyses, with a focus on ischaemic cardiomyopathy with diabetes (ICM-DM). **a**, UpSet plot summarising significant differentially expressed proteins in all the heart failure conditions; ICM-DM, ICM without diabetes (ICM-No DM), non-ischaemic cardiomyopathy with diabetes (NICM-DM), and NICM without diabetes (NICM-No DM), vs AMD from proteomic mass spectrometry (MS). Statistical significance of differential expression was determined following Benjamini-Hochberg false discovery rate adjustment (FDR) of *P* values (FDR < 0.05). **b**, Gene Ontology (GO) analysis by PANTHER (http://geneontology.org/, PANTHER17.0) enrichment bubble plot showing selected significantly enriched Biological Process (BP), Cellular Component (CC), and Molecular Function (MF) nodes from proteins which were significantly differentially expressed in all heart failure conditions vs AMD. Nodes with an FDR < 0.05 were considered statistically significant. This plot was the combination of two separate GO analyses; one from down-regulated proteins compared to AMD (blue) and one from up-regulated proteins compared to AMD (red). Enrichment count, represented as the size of the bubble, is the number of significant proteins in that particular node. GO fold enrichment is calculated as observed enrichment count/expected enrichment count from a random set of gene symbols of equal an input set size. **c**, GO analysis by PANTHER enrichment bubble plot of selected significantly enriched nodes from proteins which were significantly expressed only in ICM-DM vs AMD. **d**, UpSet plot summarising FDR significant differentially abundant metabolites in all the heart failure conditions vs AMD from metabolomic MS. **e**, Heat map of the two greatest fold change (FC) metabolites in, and specific to, ICM-DM vs AMD. **f**, Circular heatmap of all heart failure conditions log_2_ FC vs AMD quantified acylcarnitines from lipidomic MS where FDR significant differentially abundant lipids are indicated. A Lipid’s chain length was contrasted to others in colour; very long-chain (22 or more carbon length, maroon), long-chain (13-21 carbon length, orange), and medium-chain (6-12 carbon length, pink). Short-chain acylcarnitines (2-5 carbon length) were quantified in metabolomic MS.

**Supplementary Figure 6.**
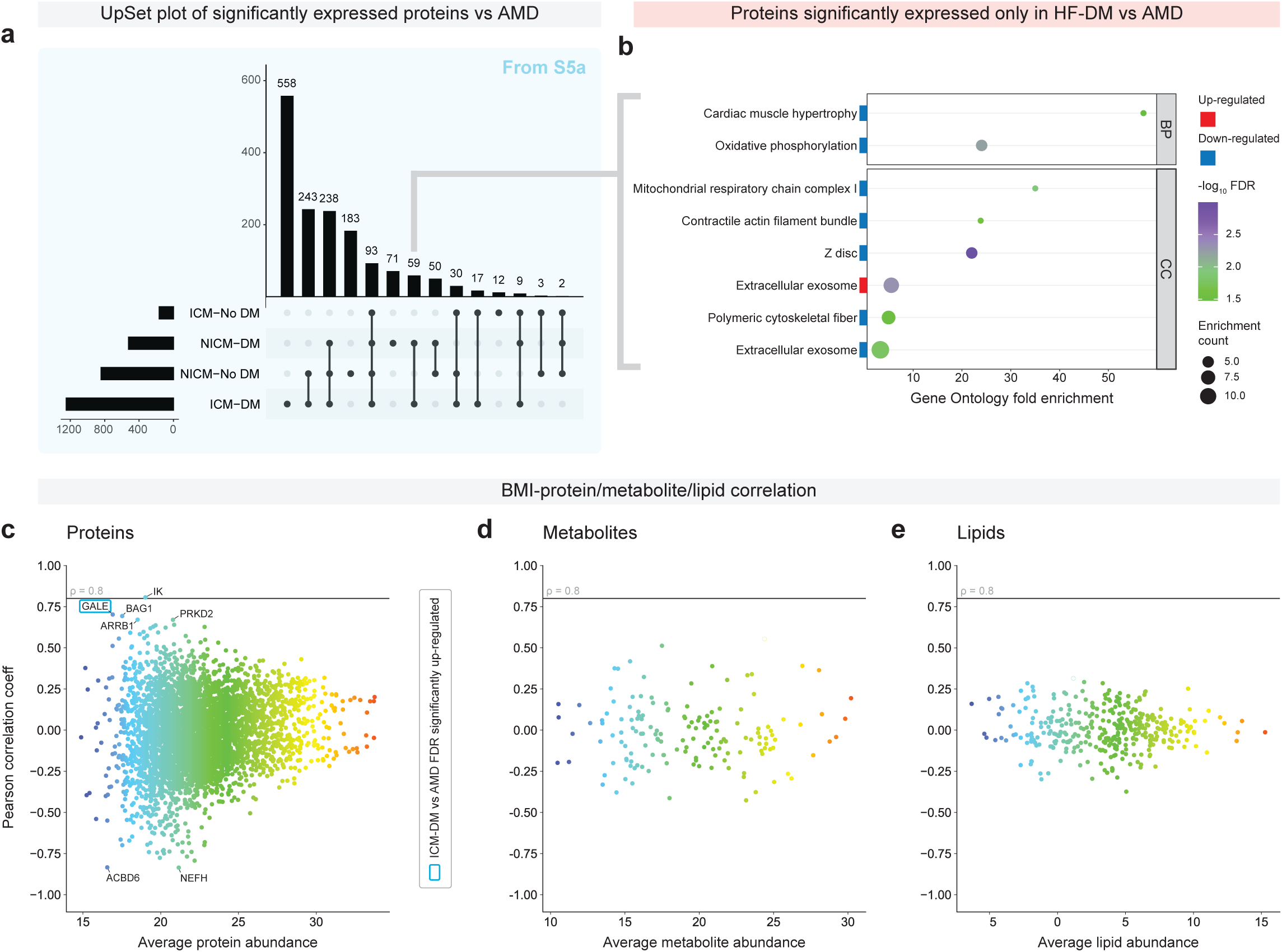
Influences of diabetes in heart failure (HF), and body mass index across healthy and HF phenotypes on myocardium molecule abundance. **a**, UpSet plot from Fig. S5a summarising significantly expressed proteins in all heart failure conditions vs age-matched donors (AMD). **b**, Gene Ontology (GO) analysis by PANTHER (http://geneontology.org/, PANTHER17.0) enrichment bubble plot showing selected significantly enriched Biological Process (BP), Cellular Component (CC), and Molecular Function (MF) nodes from proteins which were significantly differentially expressed in HF with diabetes (HF-DM; ICM-DM and NICM-DM) vs AMD. Nodes with an FDR < 0.05 were considered statistically significant. This plot was the combination of two separate GO analyses; one from down-regulated proteins compared to AMD (blue) and one from up-regulated proteins compared to AMD (red). Enrichment count, represented as the size of the bubble, is the number of significant proteins in that particular node. GO fold enrichment is calculated as observed enrichment count/expected enrichment count from a random set of gene symbols of equal an input set size. **c**-**e**, BMI-molecule (protein, metabolite, and lipid) Pearson correlation coefficient plot to determine the strength of the relationship between BMI and molecule abundance (potential additional confounder in ICM-DM HF group which has the highest BMI). Molecules with a correlation coefficient of ρ > 0.8 were accepted as being correlated. Average molecule abundance was calculated across all samples with a BMI value. Selected proteins with a high ± ρ were annotated wherein those with a blue border were also FDR significantly expressed in ICM-DM vs AMD.

**Supplementary Figure 7.**
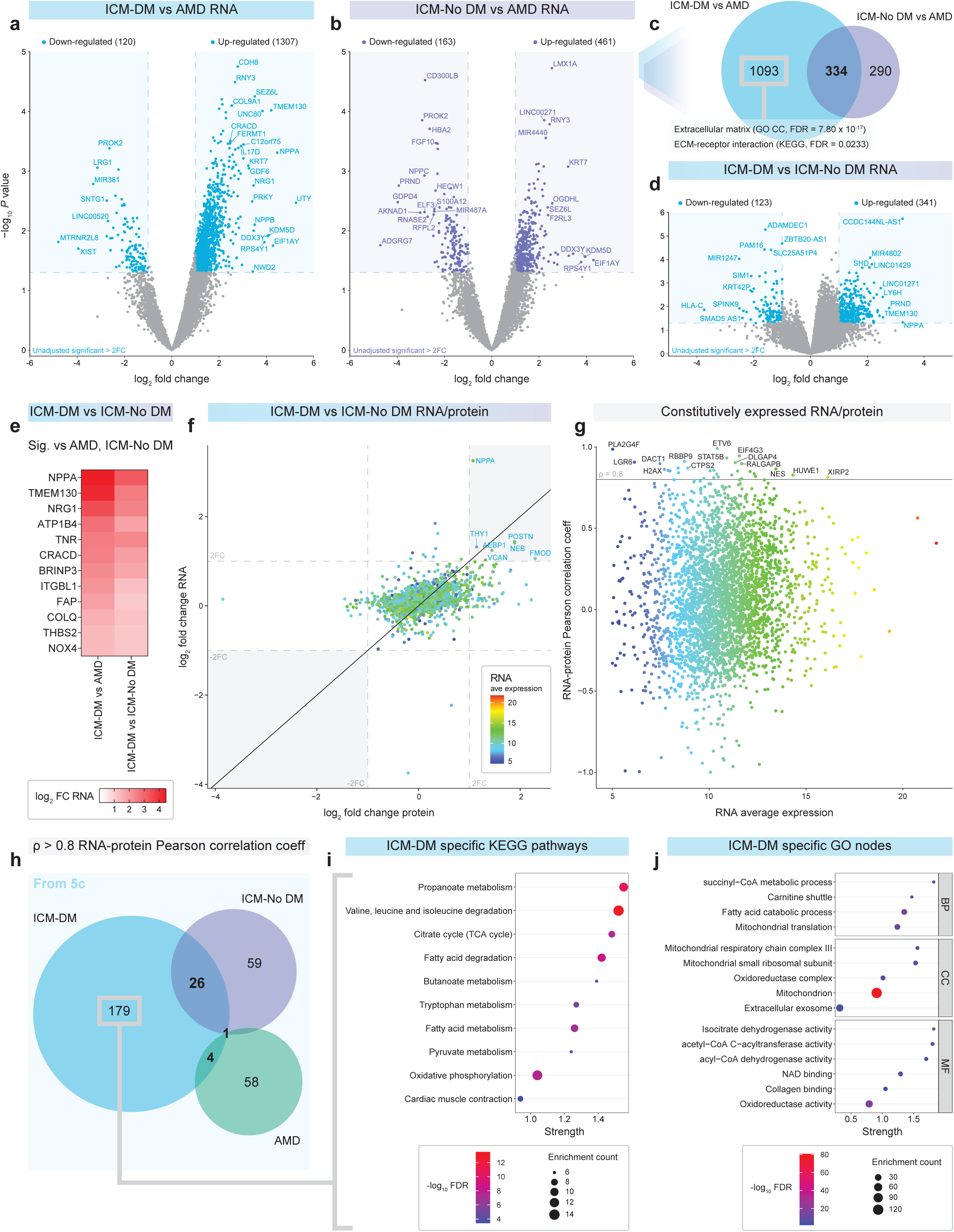
Human left ventricular myocardium RNA sequencing analyses with protein co-expression and correlation in ischaemic cardiomyopathy (ICM) and age-matched donors (AMD). **a-b**, Differential statistical significance was determined as unadjusted significant (*P* = 0.05) and > ±2-fold change (FC). Higher FC significant sequences, written as gene symbols, were annotated.. **a**, ICM with diabetes (ICM-DM) vs AMD. **b**, ICM without diabetes (ICM-No DM) vs AMD. **c**, Summary Venn diagram of Figures S7a, S7b. Sequences which were only significant in ICM-DM were analysed in STRING (https://string-db.org/, version 12.0) which revealed that the Extracellular matrix Gene Ontology (GO) Cellular Component (CC) node and the ECM-receptor interaction KEGG pathway were among the highest significantly enriched nodes/pathways. Nodes/pathways with a Benjamini-Hochberg false discovery rate adjusted *P* value (FDR) < 0.05 were determined as significant. **d**, ICM-DM vs ICM-No DM RNA. Down and up-regulation was defined as significance in ICM-DM compared to ICM-No DM. **e**, Summary heat map of RNA sequences of biological interest from Fig. S7d which were both significant in ICM-DM vs AMD and ICM-No DM. **f**, RNA and protein log_2_ FC plot of ICM-DM vs ICM-No DM where gene symbols which are > 2-FC in both protein and RNA are annotated. ICM-DM average RNA expression represented on a colour scale. **g**, RNA-protein Pearson correlation plot of ICM-DM, ICM-No DM, and AMD combined to identify common constitutively expressed RNA/protein whereby a correlation coefficient of ρ > 0.8 was accepted as being correlated. Highest correlated gene symbols were annotated. Gene symbols were spread out across the x-axis according to average RNA expression of all the groups combined. **h**, From Fig. 5c, showing gene symbols which had an RNA-protein Pearson correlation coefficient of ρ > 0.8 in ICM-DM, ICM-No DM and AMD. **i**-**j**, STRING (https://version-11-5.string-db.org/, version 11.5) analysis enrichment bubble plots of selected FDR significant (FDR < 0.05) KEGG pathways and Gene Ontology (GO) nodes from gene symbols which were RNA-protein correlated only in ICM-DM. Strength of the enriched pathways/nodes were calculated as log_10_(observed enrichment count/expected enrichment count from a random set of gene symbols of equal an input set size) wherein the enrichment count (represented as the size of the bubble) was the number of significant differentially expressed gene symbols in that particular node. **h**, Enriched KEGG pathways. **i**, Enriched GO Biological Process (BP), CC, and Molecular Function (MF) nodes.

**Supplementary Figure 8.**
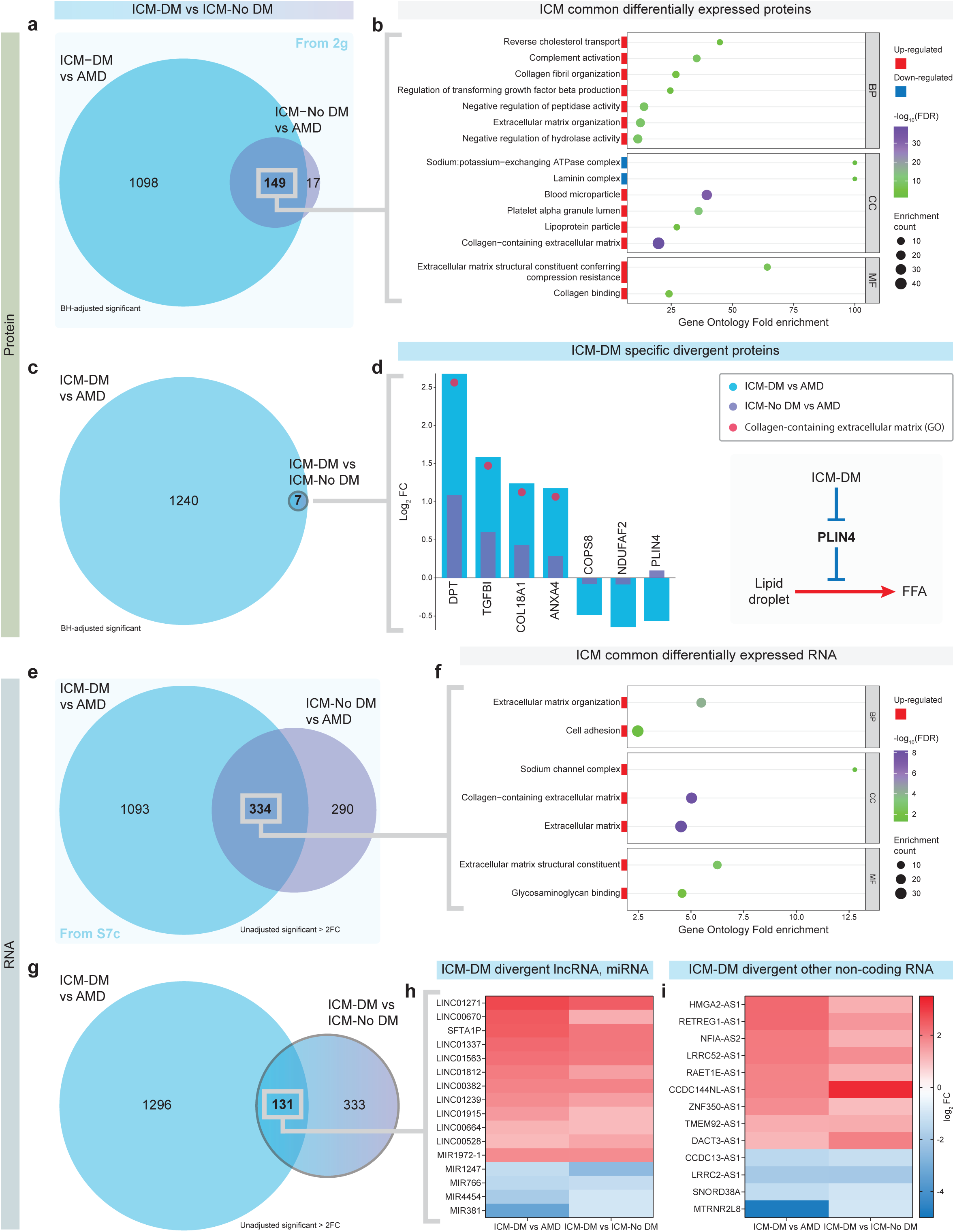
Human ischaemic cardiomyopathy (ICM) common differentially expressed myocardial proteins and RNA, and ICM with diabetes (ICM-DM) specific divergent proteins and RNA. **a**, From Fig. 2g of ICM-DM and ICM without diabetes (ICM-No DM) vs AMD significant differentially expressed proteins. Statistical significance was determined following Benjamini-Hochberg false discovery rate adjustment (FDR) of *P* values (FDR < 0.05). **b**, Gene Ontology (GO) analysis by PANTHER (http://geneontology.org/, PANTHER17.0) enrichment bubble plot showing selected significantly enriched Biological Process (BP), Cellular Component (CC), and Molecular Function (MF) nodes from proteins which were significantly differentially expressed in both ICM-DM and ICM-No DM vs AMD. Nodes with an FDR < 0.05 were considered statistically significant. This plot was the combination of two separate GO analyses; one from down-regulated proteins compared to AMD (blue) and one from up-regulated proteins compared to AMD (red). Enrichment count, represented as the size of the bubble, is the number of significant proteins in that particular node. GO fold enrichment is calculated as observed enrichment count/expected enrichment count from a random set of gene symbols of equal an input set size. **c**, ICM-DM vs AMD and ICM-DM vs ICM-No DM. **d**, A log_2_ fold change (FC) bar plot of ICM-DM and ICM-No DM vs AMD of proteins found to be significant in ICM-DM vs ICM-No DM (ICM-DM-specific and divergent from ICM-No DM). PLIN4, which was reduced in ICM-DM vs AMD while increased in ICM-No DM vs AMD, negatively regulates free fatty acid (FFA) availability. **e**, Summary Venn diagram from Fig. S7c of ICM-DM and ICM-No DM vs AMD significant differentially expressed RNA. Significance of RNA expression was determined as unadjusted significant (*P* = 0.05) and ±2-FC. **f**, GO analysis by PANTHER of significant differentially expressed RNA in both ICM-DM and ICM-No DM vs AMD. **g**, Venn diagram of significantly expressed RNA in ICM-DM vs AMD and ICM-DM vs ICM-No DM. **h**, Long non-coding and microRNA sequences, and **i**, other non-coding RNA significant in ICM-DM vs AMD and ICM-DM vs ICM-No DM in the same direction.

**Supplementary Figure 9.**
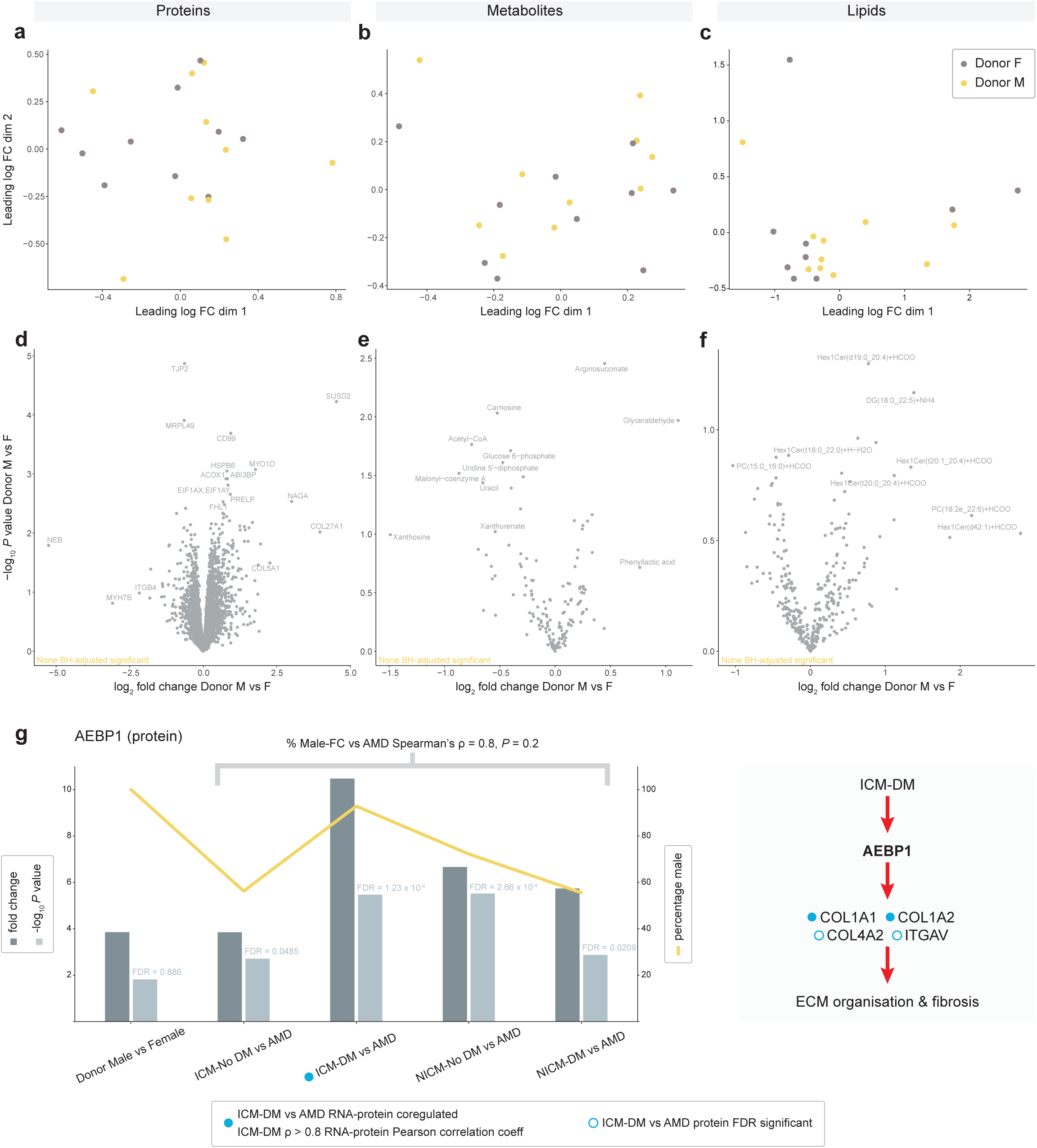
Healthy donor male myocardium vs female and potential male-dominant influence in heart failure. **a-b**, Multidimensional scaling (MDS) plots, a tool to visualise high dimensional data in two dimensions, were used to iteratively separate samples (each point) in distance based on their dissimilarity of paired data (proteins, metabolites, and lipids; dimensions) calculated as the leading log_2_ fold change (FC, average root-mean-square of the largest log_2_ FCs). Healthy donor males represented as yellow, females as grey. **d**-**f**, Volcano plots of male vs female molecule (proteins, metabolites, and lipids, respectively) abundance whereby significance was determined after Benjamini-Hochberg false discovery rate adjustment (FDR) of *P* values (FDR < 0.05). The was no significance in abundance between donor males and females. **g**, Bar plot of protein AEBP1 fold change (dark grey) and log_10_ *P* value (light grey, with FDR adjusted value annotated) in donor males vs females, and heart failure conditions vs age-matched donors (AMD). Superimposed line plot shows the associated percentage of males in the subject group; the donor male group are 100% male. AEBP1 was FDR significant differentially expressed in all HF conditions vs AMD but the most in ICM-DM, which also had the highest FC. The ICM-DM group had the highest percentage of males. A male-protein FC vs AMD nonparametric Spearman’s correlation test was performed on the HF conditions to determine a possible significant male relationship to AEBP1 FC. Significance was determined as ρ > 0.8 and *P* < 0.05. AEBP1 has been identified to positively regulate extracellular matrix (ECM) organisation and fibrosis, and was co-regulated in ICM-DM vs AMD in both RNA and protein, and RNA-protein correlated in ICM-DM.

**Supplementary Figure 10.**
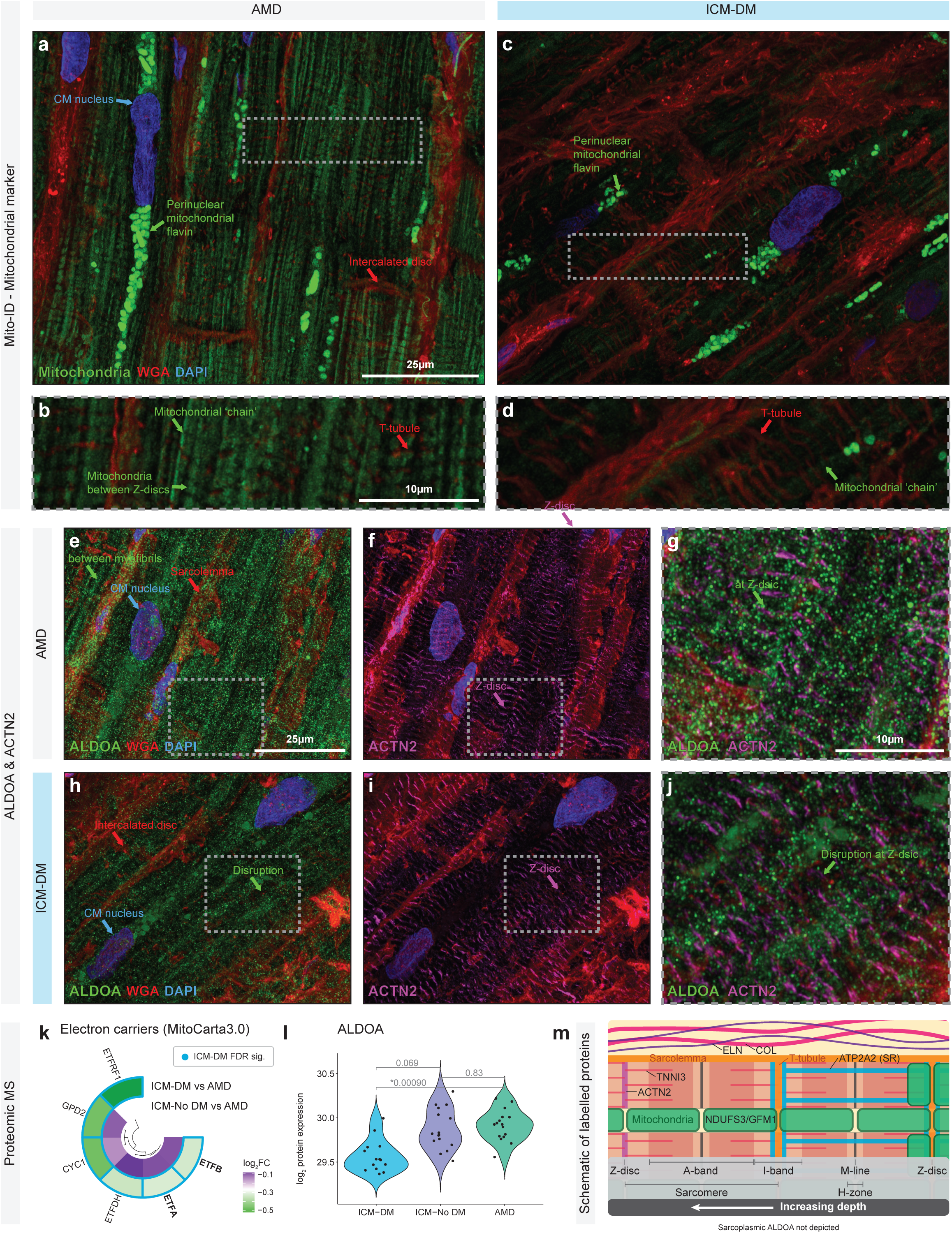
Immunofluorescent labelling of mitochondria and sarcoplasmic ALDOA in cryopreserved ischaemic cardiomyopathy with diabetes (ICM-DM) and healthy age-matched donor (AMD) myocardium. **a-b**, Immunofluorescent confocal microscopy images of cryopreserved human myocardium representing qualitative differences between AMD and ICM-DM in mitochondria labelled with Mito-ID. Bright (autofluorescent) perinuclear mitochondrial flavin identified. **e**-**j**, Labelled muscle aldolase (ALDOA, green), localised in the sarcoplasm, with a labelled Z-disc protein, ACTN2 (magenta). Membranes, particularly the sarcolemma, were stained with fluorophore-conjugated wheat germ agglutinin (red) and nuclei were stained using DAPI (blue). Cardiomyocytes (CM) are depicted in a longitudinal orientation. All images are 4.5µm thick Z-stacks, deconvolved using Huygens Professional, and compressed into a two-dimensional image using Fiji/ImageJ Maximum Intensity Projections. **k**, Proteomic mass spectrometry (MS) log_2_FC circular heatmap of quantified proteins from the MitoCarta3.0 electron carriers gene set. **l**, Violin plot of proteomic MS log_2_ transformed quantification of ALDOA in ICM-DM, ICM without diabetes (ICM-No DM), and AMD groups. FDR adjusted *P* values were annotated to reveal significant differences (*, FDR < 0.05) between groups. **m**, Cellular schematic showing localisation of all histologically labelled proteins in this study. Sarcoplasmic ALDOA not depicted. SR; sarcoplasmic reticulum. All tissue was pre-mortem. Macroscopic scar tissue, particularly in heart failure conditions, was avoided in all quantitative analyses (mass spectrometry and RNA sequencing) and imaging. Only the most normal/healthy appearing and longitudinally oriented AMD and ICM-DM cardiomyocytes were imaged in confocal microscopy.

**Supplementary Figure 11.**
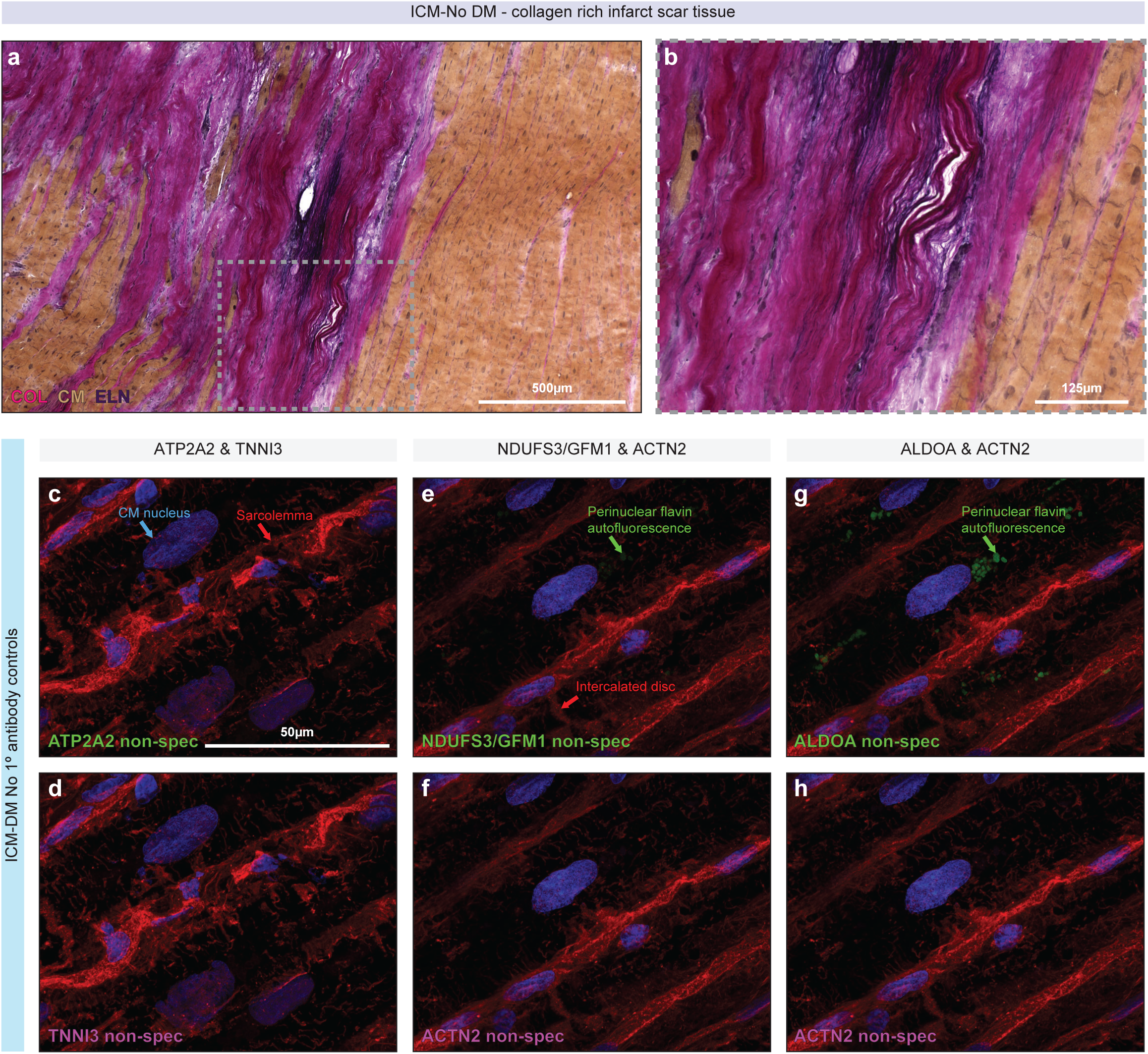
Brightfield microscopy example of avoided macroscopic myocardial scar tissue and immunofluorescent microscopy control images. **a**-**b**, Bright-field microscopy images of Verhoeff-Van Gieson stain of ischaemic cardiomyopathy without diabetes (ICM-No DM) macroscopic infarct/replacement fibrosis scar tissue with an inset at a higher magnification. Collagens (COL), elastin (ELN, dark purple), cardiomyocytes (CM, brown), and nuclei (dark blue) identifiable. Macroscopic scar tissue in all heart failure tissues was avoided for all quantitative analyses (mass spectrometry and RNA sequencing) as well as in all microscopy. **c**-**h**, ICM-DM no primary antibody control stains to detect any autofluorescence/non-specific fluorophore-conjugated secondary antibody labelling within the myocardium. Columns are in reference to the sections co-stained with that combination of primary antibodies shown in Figures 6d-e and S10e-j. All paired sections (AMD and ICM-DM) were cut (16µm at -16°C), co-stained, imaged, and post-processed under identical conditions at the same time; antibodies and fluorophore conjugated stains were from the same master mix, sections were imaged under identical settings, and brightness/contrast adjusted identically in Fiji/ImageJ post-processing. All fluorophore conjugated secondary antibodies were applied with a concentration of 5µg/mL to all sections. NDUFS3, GFM1 and ALDOA featured stains were stained in the same batch, so were referenced to the same no primary antibody control section. Experimental condition (ICM-DM) was chosen for no primary antibody control. Membranes, particularly the sarcolemma, were stained with fluorophore-conjugated wheat germ agglutinin (red) and nuclei were stained using DAPI (blue). All images are 4.5µm thick Z-stacks, deconvolved using Huygens Professional, and compressed into a two-dimensional image using Fiji/ImageJ Maximum Intensity Projections. All tissue pre-mortem.

## Notes

### Competing Interest Statement

The authors have declared no competing interest.

### Clinical Trial

NA

### Author Declarations

The methods of procurement, storage and use of donated human myocardium were approved by the Human Research Ethics Committee at The University of Sydney (USYD 2021/122).

